# First-in-human demonstration of splenic ultrasound stimulation for non-invasively controlling inflammation

**DOI:** 10.1101/2020.07.14.20153528

**Authors:** Rachel S. Graham, Daniel P. Zachs, Victoria Cotero, Catherine D’Agostino, Despoina Ntiloudi, Claire R.W. Kaiser, John Graf, Kirk Wallace, Richard Ramdeo, Thomas R. Coleman, Jeffrey Ashe, John Pellerito, Kevin J. Tracey, Bryce A. Binstadt, Sangeeta S. Chavan, Stavros Zanos, Christopher Puleo, Erik Peterson, Hubert H. Lim

## Abstract

Hyperinflammation and uncontrolled cytokine release in infections and autoimmune diseases require therapy to reduce the innate immune response. Here, we present first in-human data showing reduction in pro-inflammatory cytokine release with ultrasound stimulation of the spleen in healthy subjects and in rheumatoid arthritis patients. Single cell RNA sequencing reveals a decrease in IL-1β and IL-8 transcript levels in circulating monocytes. There is also a down regulation of pathways involved in TNF and IL-6 production, and IFNγ- and NFκB-regulated genes. Additional pre-clinical studies reveal that ultrasound can boost B cell activation and antibody production. Splenic ultrasound offers a new non-invasive therapy for treating hyperinflammation without compromising the adaptive immune response.

## Introduction

Cytokine release by cells of the innate immune system drive inflammation (*1, 2*). Inflammatory reactions are typical in response to microbial or viral infection but can lead to health problems or life-threatening conditions if there is a persistent hyperactive innate immune response, involving cytokine toxicity and tissue damage (*2, 3*). In addition to other acute conditions, such as sepsis and acute kidney injury (*4, 5*), hyperinflammation is an issue that occurs across multiple chronic inflammatory systemic diseases, such as rheumatoid arthritis (RA) and irritable bowel syndrome (*6, 7*). Current pharmacologic or biologic approaches to treating hyperinflammation are associated with multiple side effects and high costs.

Over the past 20 years, researchers within the field of bioelectronic medicine have investigated an unconventional approach for treating inflammation through the use of vagus nerve stimulation (*8-10*). Electrical stimulation of the vagus nerve activates the splenic nerve and cells within the spleen, leading to activation of the cholinergic anti-inflammatory pathway (*11, 12*). This pathway depends on splenic nerve release of norepinephrine that activates acetyltransferase - expressing T lymphocytes, which in turn modulates innate immune cells to decrease systemic levels of key pro-inflammatory cytokines, such as IL-6, IL-15 and TNF (*13, 14*). Furthermore, electrical vagus nerve stimulation has been shown in mice and humans to provide therapeutic anti-inflammatory effects for chronic inflammatory diseases (e.g., RA and irritable bowel syndrome) and for acute inflammation (e.g., sepsis, renal ischemia, trauma/hemorrhagic shock, and acute lung injury following trauma/hemorrhagic shock; (*13, 15-19*)).

Direct vagus nerve stimulation requires invasive implantation procedures. Several groups have pursued non-invasive ultrasound stimulation of the spleen as a safer, non-surgical approach for modulating the cholinergic anti-inflammatory pathway. Since the vagus nerve projects to the brain and multiple organs throughout the body (*20*), targeting neurons or cells specifically within the spleen can reduce unintended activation or side effects. There has been a recent surge of research that demonstrates the ability to activate or modulate cells with ultrasound energy (*21-26*). There are several studies in rodents showing the ability to modulate the splenic nerve or immune cells within the spleen with ultrasound, to reduce inflammation and cytokine levels (*27-31*). One of the initial reports was in a mouse model of reperfusion injury and kidney inflammation, where ultrasound stimulation of the spleen significantly reduced kidney damage (*27, 32*). Recently, our research groups discovered that specific parameters of ultrasound stimulation of the spleen can drive significant anti-inflammatory effects in rodent models of both acute and chronic inflammation (i.e., sepsis and inflammatory arthritis; (*29, 30*)), which was shown to be mediated through a similar cholinergic anti-inflammatory pathway accessed through vagus nerve stimulation. Ultrasound is a potentially impactful clinical solution to inflammation as it can be applied non-invasively to the body with a wearable device and with energy parameters already shown to be safe for the human body based on numerous ultrasound imaging applications (*33, 34*).

Here, we show the first in-human results of pro-inflammatory cytokine reduction with non-invasive ultrasound stimulation of the spleen. In healthy individuals, a single three-minute administration of splenic ultrasound stimulation significantly inhibits whole blood TNF production upon *ex vivo* exposure to endotoxin compared to sham controls. In RA patients, we observed that daily splenic ultrasound stimulation results in reduction of blood-borne transcripts encoding for pro-inflammatory markers IL-1β, IL-8, and NFκB, as well as suppressing pathways involved in IL-6 and TNF production. Ultrasound also reduces pathways involved with monocyte migration, which may contribute to its anti-inflammatory effect. From a safety perspective, circulating immune cell composition does not change with ultrasound treatment, and ultrasound does not inhibit the adaptive immune response in humans. Our additional pre-clinical animal data further demonstrates that splenic ultrasound stimulation results in an enhanced antibody response upon exposure to an inflammatory antigen. Thus, activation of the splenic neuroimmune pathway may provide a low risk therapeutic approach for a broad range of health conditions, due to its pleiotropic nature and physiological role in suppressing specific cytokines involved in innate immunity while enhancing the transition to and maintenance of the adaptive immune response (*15, 35, 36*). Together, these data support a new non-invasive clinical therapy via splenic ultrasound stimulation for a range of acute or chronic inflammatory diseases without requiring surgery or intake of artificial substances with possible side effects.

## Results

### Non-invasive Splenic Ultrasound Reduces TNF in Humans

To investigate how non-invasive ultrasound stimulation of the spleen affects cytokine levels directly in human subjects, healthy participants were recruited into a study designed to investigate the effects of ultrasound stimulation in the spleen on cytokine response to ex vivo endotoxin exposure. The first immunomodulation results from an ongoing feasibility clinical study are presented here; additional investigations on the effects of different ultrasound stimulus placement and the effects of altering the applied ultrasound pressure will be presented in subsequent manuscripts (feasibility study described at clinicaltrials.gov: NCT03548116).

Healthy participants received pulsed ultrasound stimulation of the spleen for three minutes (2.2 MHz, 1 Hz pulse repetition frequency, 290.4 mW/cm^2^ Ispta) or sham stimulation, which followed the same procedure with no ultrasound energy being delivered to the spleen. Changes in inflammatory response status were then tested using whole blood collected from the subjects following the ultrasound (or sham) treatment and challenged with lipopolysaccharide (LPS) *ex* vivo. This *ex vivo* cytokine production assay has been previously used to verify splenic neuroimmune activation using implanted vagus nerve stimulators and is a well-established method for non-invasively assessing activation of neuroimmune pathways (*15, 37-39*). LPS-induced TNF production in blood samples from ultrasound or sham stimulated subjects was tested at multiple LPS concentrations (Fig. 1A); a significant reduction in TNF was observed in the ultrasound stimulated group compared to the non-stimulated group (Fig. 1B; p-value = 0.006 using Wilcoxon unpaired rank-sum test) at the optimal LPS concentration (1 ng/mL; Fig.1A). These data are consistent with earlier reports using implanted nerve stimulators or pharmaceutical activators of the splenic pathway (*15, 38*). They are also consistent with the magnitude of TNF reduction using similar ultrasound stimulation parameters in a previous study activating the neuroimmune pathway in a rodent model of using Wilcoxon unpaired rank-sum test) at the optimal LPS concentration (1 ng/mL; Fig.1A). These data are consistent with earlier reports using implanted nerve stimulators or pharmaceutical activators of the splenic pathway (*15, 38*). They are also consistent with the magnitude of TNF reduction using similar ultrasound stimulation parameters in a previous study activating the neuroimmune pathway in a rodent model of endotoxemia (*29*). Further supporting the human results, we also demonstrated that splenic ultrasound stimulation in a rodent model achieves a reduction across a number of pro-inflammatory cytokines beyond TNF, including IL-6, IFNγ, IL-1β, IL- 1α, and IL-12 in splenic lysates (Fig. S1), which is consistent with previous reports using implanted vagus nerve stimulators (*15, 40*).

**Figure 1.**
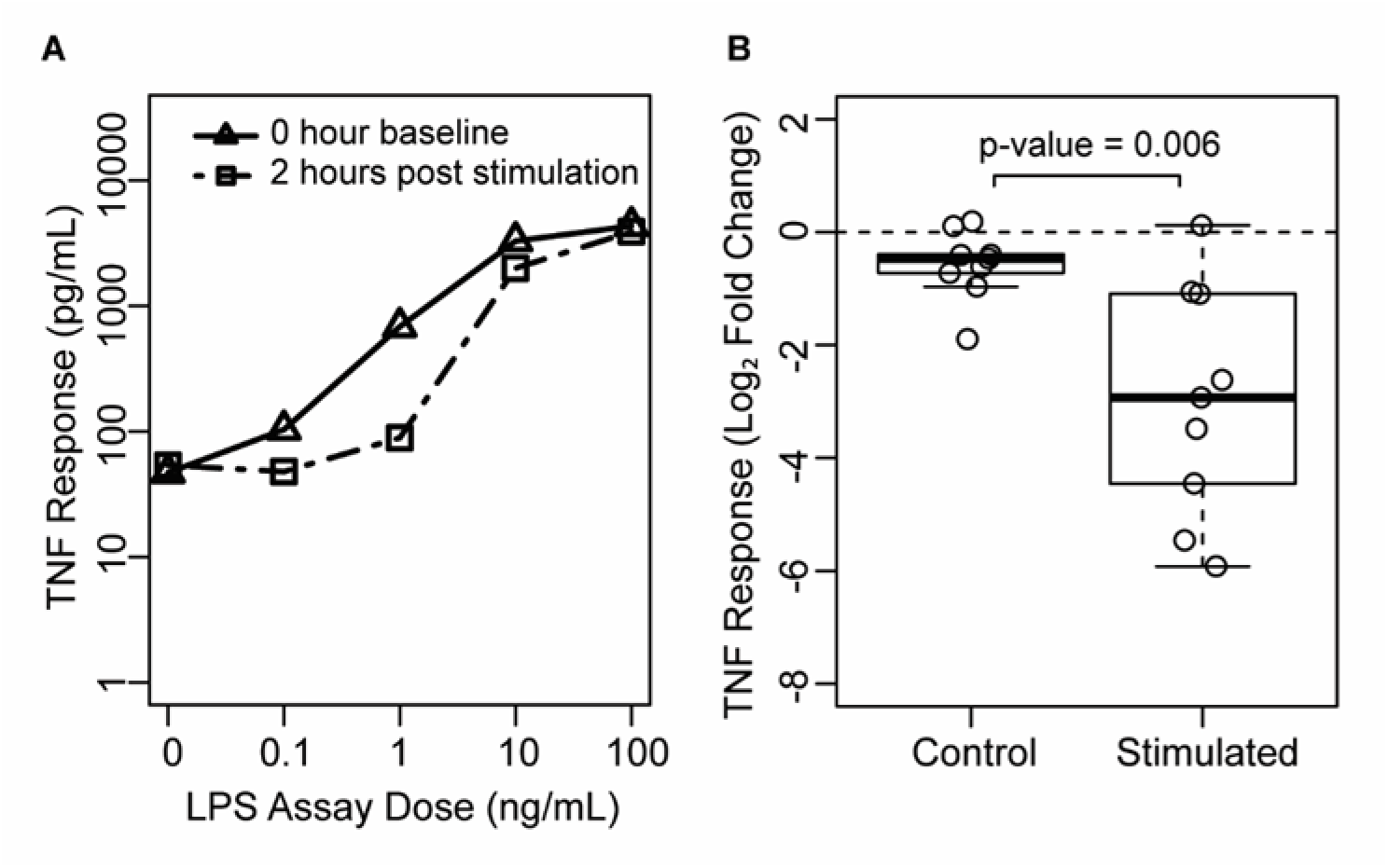
Non-invasive ultrasound stimulation of the spleen reduces TNF production in human subjects. A) Whole blood from all subjects and samples was incubated with LPS at multiple concentrations. TNF response after LPS incubation is shown for one ultrasound stimulated subject from samples collected before (0 hours baseline) and after (2 hours post stimulation) ultrasound application. The optimal dose of LPS for measuring the immunomodulatory response to ultrasound was found to be at 1 ng/mL, and this is used for comparing the TNF response between the ultrasound stimulated versus control groups. B) Ultrasound stimulation of the spleen decreases whole-blood LPS-induced TNF release, where blood was obtained from 18 healthy human subjects prior to and then two hours following splenic ultrasound application (stimulated: n=9, control: n=9). The data shown were taken from the 1 ng/ml LPS concentration for each dose response curve for all subjects and samples. *P*-value was computed using the unpaired Wilcoxon rank- sum test.

### Pro-inflammatory Cytokines and Signaling Pathways are Downregulated in Circulating Monocytes with Ultrasound Treatment

Previous animal research demonstrated that ultrasound can significantly reduce inflammation and improve clinical measures in a chronic inflammatory arthritis model (*30*). Expanding upon the findings in healthy human subjects in the previous section, we present data from a clinical study consisting of splenic ultrasound treatment in RA patients (clinical study described at clinicaltrials.gov: NCT03690466). Participants received ultrasound stimulation of the spleen with the goal of lowering the over-active inflammatory response that occurs during RA flare-ups. PBMCs were isolated from whole blood taken from each research participant before ultrasound treatment (day 0, pre-stimulation) and after two weeks of daily 30-minute sessions of ultrasound to the spleen (day 14, post-stimulation). From these 10 samples (subjects = 5, timepoints = 2), a total of 53,343 PBMCs were successfully sequenced from single cells (Table S1). *In silico* analysis identified 13 cell-types based on gene expression patterns of standardly used marker genes (Fig. S2). Participants received stimulation with a 1 MHz transducer (1.2 W/cm^2^; SoundCare Plus, Roscoe Medical) that was moved continuously across a 5-inch by 5-inch square, centered on the spleen, as identified by an ultrasonographer.

**Figure 2.**
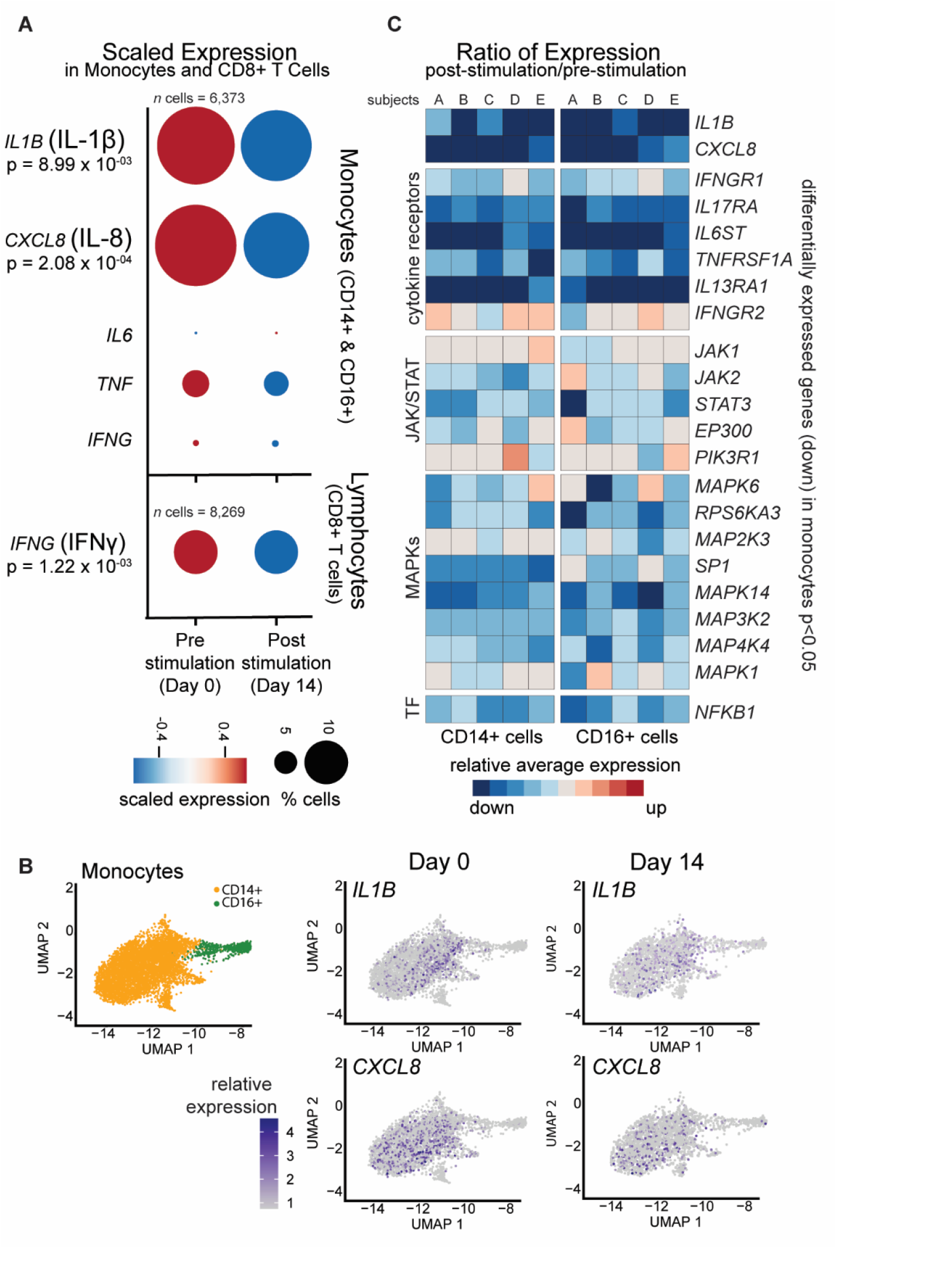
Splenic ultrasound treatment reduces cytokines and downstream pathways in-vivo in RA patients. A) Single cell RNA sequencing of PBMCs from RA patients receiving 14 days of 30-minute daily ultrasound stimulation of the spleen. Samples were collected pre- (day 0) and post-stimulation (day 14) with ultrasound. Dot plot shows percent of CD 14+ and CD 16+ monocytes (top) or CD8+ T cells (bottom) that express the transcript (labeled on the left) represented by size and scaled expression level represented by color (obtained by subtracting the mean and dividing by the standard deviation). Key pro-inflammatory cytokines are shown with unadjusted p-values labeled for those transcripts significantly downregulated (p-value <0.05). B) UMAP showing top two dimensions (principal components) contributing to variability within the dataset. UMAP diagrams showing CD 14+ and CD 16+ monocytes represented by orange and green dots, respectively. IL-1|3 and CXCL8 expression on day 0 and day 14 are alsc mapped on these UMAP diagrams. C) Heatmap displaying the average relativ expression (for all cells in specified group) of downregulated transcripts (genes listed on the right, p-value <0.0: Color denotes relative expression for post-ultrasound levels compared to (divided by) pre-ultrasound levels for each subject (represented by columns, i = 5, participant A-E shown left to right in CD14+ cells or CD16+ cells. Genes encoding for cytokine receptors, components of the JAK/STAT pathway MAP kinases, and transcription factor (TF) are grouped and labeled on the left of the plot.

We aimed to identify molecular changes that occur with splenic ultrasound in humans based on single cell RNA sequencing to better understand how this ultrasound treatment can drive an anti-inflammatory response. We observed reduced monocyte expression of key pro-inflammatory cytokine genes encoding for IL-1β and IL-8 from all five subjects post-stimulation compared with pre-stimulation (Fig. 2A, p-values given for transcripts showing significance of p<0.05). Although this result was determined by analyzing all monocytes, reduction of these two markers appears primarily in CD14+ monocytes as this is the cell type showing predominant expression of those genes (Fig. 2B). The same analysis in CD8+ T cells show a decrease in transcripts that encode for IFNγ (Fig. 2A). The changes in transcript level of these cytokines can only be detected when comparing within the same cell type before and after treatment, within the cell that expresses the cytokine (Fig. S2). Consistent with known expression patterns of *IL1B* and *CXCL8* transcripts (encoding for IL-1β and IL-8), we see that these transcripts are only suppressed in the monocytes where they are primarily expressed. Likewise, *IFNG* transcripts (encoding for IFNγ) are detected and found to be reduced in CD8+ T cells (Fig. S2). In our dataset, other cytokines, such as TNF and IL-6, did not show a significant change in monocytes; however, IL-6 transcripts were detected in only 66 of the 53,343 cells. Therefore, we cannot definitively conclude if there is a change in transcript levels for those molecules due to detection limitations. Also it is possible that pro-inflammatory cytokines (such as TNF) are differentially regulated through post-transcriptional production after ultrasound treatment (*41*).

Since monocytes play a critical role in the innate immune system and the inflammatory response, we further focused our analyses on transcriptional changes that occur in circulating monocytes after ultrasound treatment. Differential expression analysis of all monocytes across the five subjects, pre- and post-ultrasound stimulation, shows 938 differentially expressed genes (p-value<0.05) with 841 of those genes being down-regulated. Additionally, this reduction in transcript levels did not appear as a result of sequencing differences, as we observe no obvious difference in the number of cells sequenced, the number of sequencing reads, or number of average genes detected per cell between monocytes taken from day 0 as compared to day 14 (Table S1, ‘CD14 Mono’ and ‘CD16 Mono’). Although these gene transcripts are defined as differentially expressed when comparing monocytes from all subjects (Fig. 2A), we also observe that for most genes, this down-regulation is consistent across subjects (Fig. 2C; columns represent cell-type averages for CD14+ and CD16+ cells for each subject that are similar across columns for many of the genes). These results are quite constant despite patient heterogeneity in age, sex, physical characteristics, disease properties and past treatments across subjects (Participants A-E in Table S2 correspond to columns 1-5 in Fig. 2C, respectively). Among down-regulated genes in monocytes, we also observe that many cytokine receptors, such as those for TNF, IL-6, IL-17, IL-13, and IFNγ are reduced with ultrasound treatment (Fig. 2C). Furthermore, components of signaling pathways downstream of these cytokine receptors are down regulated, including components of the JAK/STAT pathway, MAP kinases, and pro-inflammatory transcription factor NFκB (Fig. 2C). Together, these findings demonstrate a consistent and robust reduction in transcripts encoding key pro-inflammatory cytokines and cellular signaling molecules downstream of their receptors after 14 days of daily splenic ultrasound treatment in humans with active rheumatoid arthritis.

### Suppression of Multiple Pathways that Contribute to the Pro-inflammatory Response

Cytokines trigger multiple cellular pathways, which then contribute to systemic inflammation. Our transcriptomic data reveal that many genes known to be regulated by IFNγ and NFκB are subsequently downregulated with ultrasound treatment, further promoting a cascade of suppression across multiple pro-inflammatory pathways with ultrasound treatment (Fig. 3A). We sought to determine if the 840 down-regulated genes in monocytes showed statistical enrichment for functional Gene Ontology (GO) terms associated with inflammation. The GO system classifies genes based on their functional characteristics. Gene sets can then be analyzed for GO terms that are statistically over-represented among that set, allowing for a better understanding of which biological processes (e.g., inflammation) may be modulated with treatment. Out of the top statistically enriched GO terms for the down- regulated genes, we observe many that contribute to pro- inflammatory cellular processes. Genes suppressed by ultrasound show enrichment of GO terms including “inflammatory response” (p-value = 1.50 E-5, Table S3), “cytokine-mediated signaling pathway” (p- value = 9.7 E-5, Fig. 3B), and “cellular response to cytokine stimulus” (p- value = 6.7 E-6, Fig. 3B). We also observe enriched GO terms associated with positive regulation of TNF, IL-6, and IL-8 among the down-regulated genes, suggesting that inducing signals for TNF or IL-6 are also suppressed, despite the absences of significant change in TNF or IL-6 transcript levels (Fig. 3B). These GO term results show that many gene transcripts encoding for cellular inflammatory functions are reduced after ultrasound treatment.

**Figure 3.**
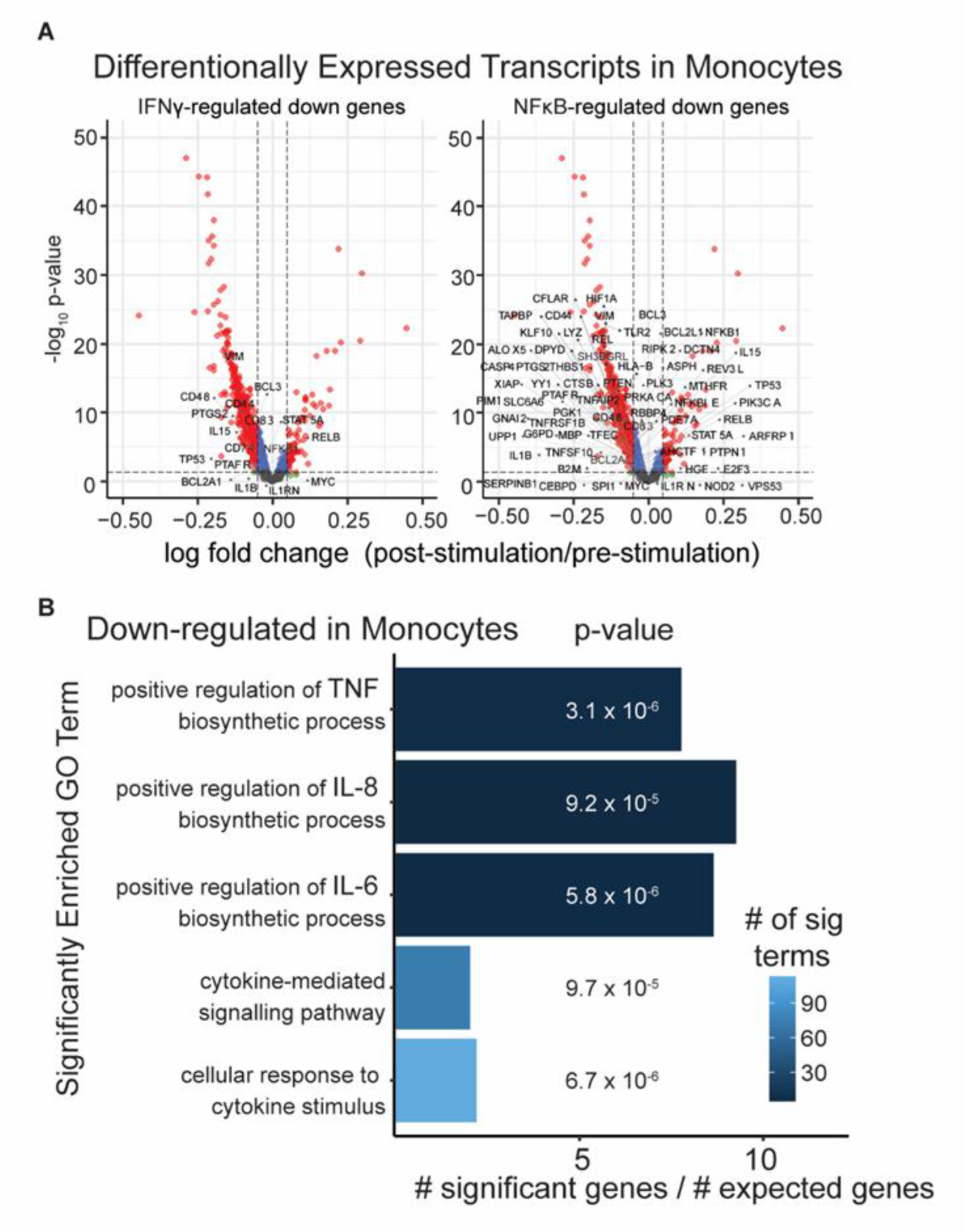
Reduction of pro-inflammatory pathway components in monocytes with ultrasound treatment. A) Volcano plot showing all detected genes in monocytes, highlighting differentially expressed genes (p-value <0.05) and labeled IFNγ-regulated genes that are down regulated, as well as NFκB-regulated genes that are down regulated as characterized by KEGG pathway gene lists. B) Gene ontology (GO) analysis using Bioconductor package ‘topGO’, determining functional GO terms significantly enriched within downregulated genes in monocytes. Enriched GO terms associated with cytokine production are shown, where bars represent number of significant genes in a list relative to number of genes that would be expected by chance, and with number of genes contributing to each GO term represented by a blue color. P-values for each term shown is from Fisher exact test.

### Down-regulation of Components Involved in Monocyte Migration

Within circulating monocytes, we observe ultrasound-treatment associated GO term enrichment of cellular components associated with IL-8 functionality (Fig. 3B), as well as a robust reduction in IL-8 across subjects (Fig. 4A, columns represent subject cell type averages). IL-8 is a chemokine (chemotactic factor) that stimulates neutrophil movement towards inflammatory sites. Therefore, we further assessed if ultrasound influences other signaling molecules involved with cellular migration (e.g., to sites of inflammation). GO term analysis of down-regulated genes in CD14+ monocytes show enrichment of “positive regulation of cell migration” among the top 15 significantly enriched GO terms. Other GO terms, such as “lamellipodium assembly” (facilitates cell mobility), “ephrin receptor signaling” (promotes cell adhesion), and “actin nucleation” (involved with cell locomotion), all suggest that ultrasound treatment induces down regulation of genes involved with cell migration (Fig. 4B). Furthermore, genes that more specifically contribute to “monocyte chemotaxis” are down-regulated post-ultrasound stimulation in CD14+ monocytes and many of these genes are consistently reduced across all five subjects (Fig. 4A). Together these observations suggest that splenic ultrasound suppresses molecular correlates of monocyte migratory capacity in humans with rheumatoid arthritis.

**Figure 4.**
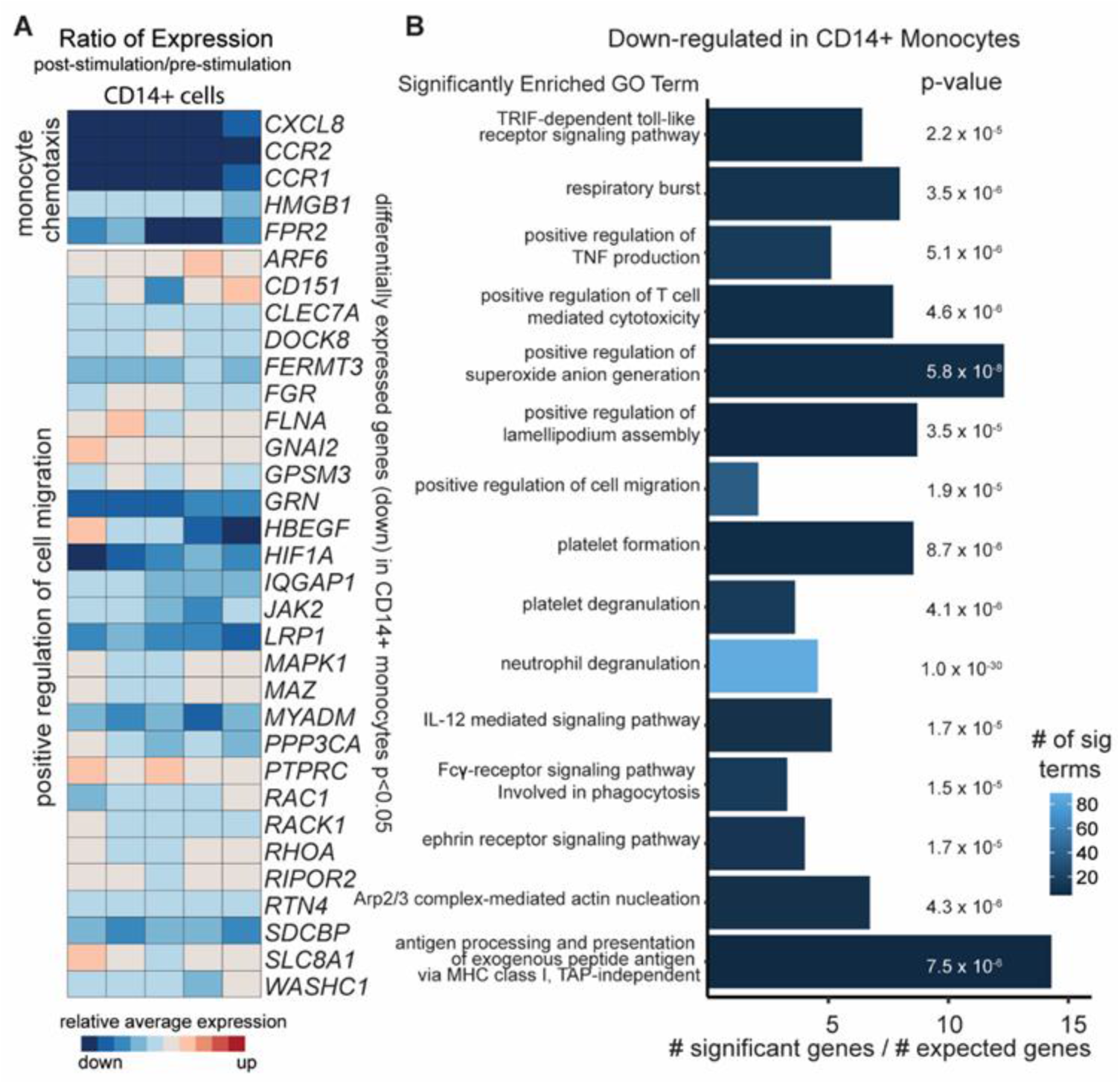
Ultrasound results in downregulation of cellular migration components in CD14+ monocytes. A) Heatmap displaying the average relative expression (for all cells in specified group) of significantly downregulated transcripts (genes listed on the right) for post-ultrasound compared to (divided by) pre-ultrasound levels for each subject (represented by columns, n = 5) in CD14+ cells. Genes encoding components involved in monocyte chemotaxis and cell migration are grouped and labeled to the left. B) Enriched GO terms within downregulated genes in CD14+ cells are shown, where bars represent the number of significant genes in the list relative to the number of genes that would be expected by chance, and with the number of genes contributing to each term represented by blue color. *P*-value for each term shown is based on Fisher exact test.

### Immune Cell Repertoire Unchanged with Moderate Degree of Suppression

We sought to determine how ultrasound affects the circulating immune cell repertoire. In comparing the transcriptional profiles (based on principal component analysis and UMAP dimensional reduction) of all circulating peripheral blood mononuclear cells pre- and post-ultrasound stimulation, we observed remarkable consistency in the composition of the celltype repertoire across subjects (Fig. 5A). Also, the calculated relative percent of each cell-type for each subject is displayed, and based on a paired Wilcoxon rank sum test for pre- and post-stimulation for each cell type, we observe no significant difference in the relative proportions (percent contribution) of the 13 cell types (Fig. 5B, statistics shown in Table S4). Additionally, when comparing the number of cells sequenced for each cell type per participant, there is no significant difference between timepoints (i.e., no technical bias, Table S4).

**Figure 5.**
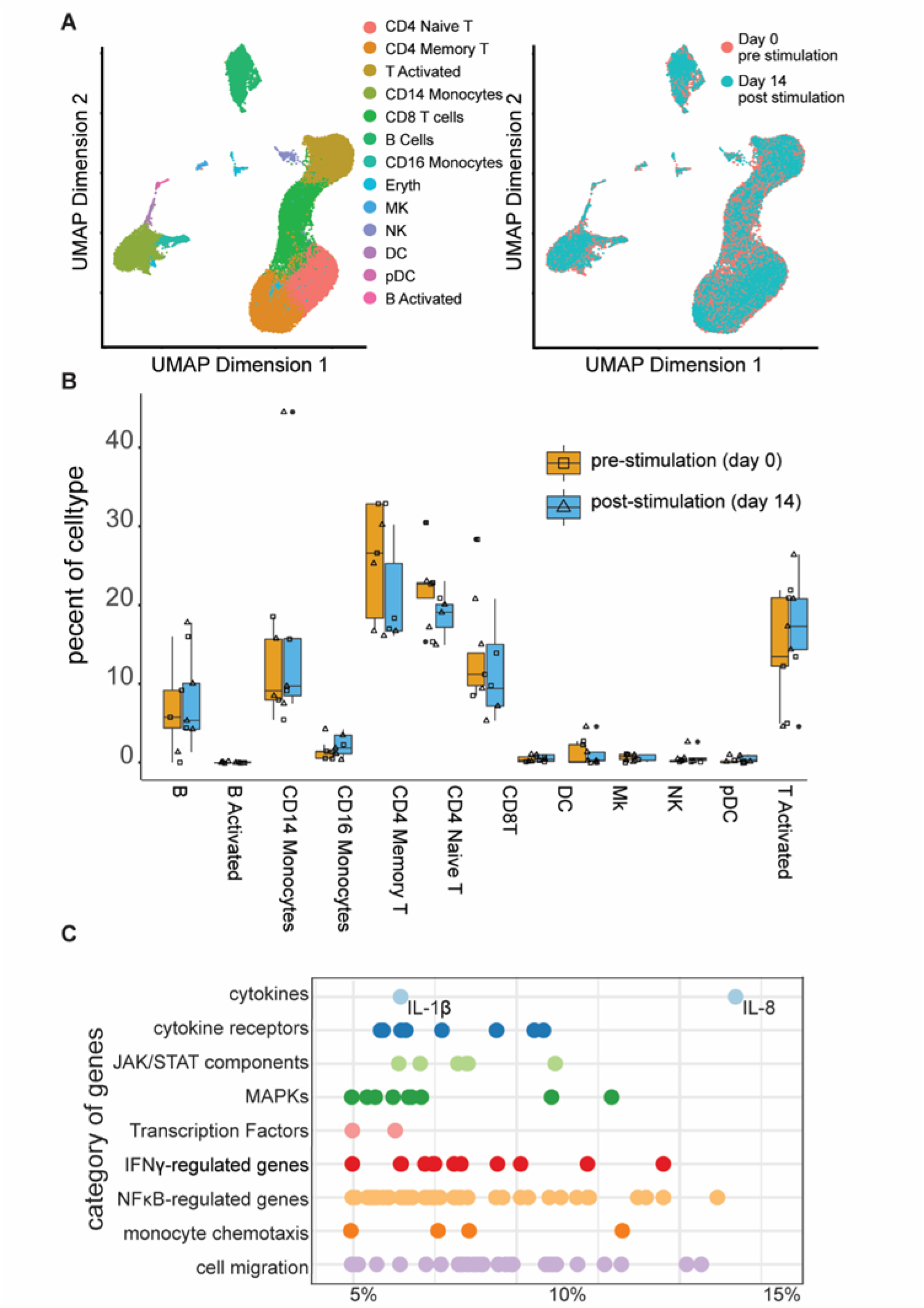
Ultrasound provides a moderate effect on the immune system. A) UMAP showing top two dimensions (principal components) contributing to variability within the dataset. All cells are shown, consisting of 53,343 cells from 10 samples (subjects = 5, timepoints = 2) with cell type and timepoint represented by color showing transcriptional signatures and relative number of cells is consistent between timepoints. B) Box plot showing median percent of each cell type (subjects, n =5) captured before ultrasound stimulation (day 0, orange, open squares) and after ultrasound stimulation (day 14, blue, open triangles) with filled in circles representing outliers. No statistical difference was detected between timepoints for any cell type based on a paired Wilcoxon ranked-sum test (Table S4). C) Percent down regulation range is shown after 14 days of ultrasound stimulation for each gene (represented by dots) in each functional category described previously.

We further determined what degree of suppression occurs after ultrasound stimulation for genes involved in inflammation (genes shown in Fig. 2C, 3A and 4A). Most differentially- expressed genes show a ~5-15% reduction in transcript levels after ultrasound treatment, suggesting a modest degree of suppression, with IL-8 and genes involved in cell migration showing the highest degree of suppression (>15%, Fig. 5C). These findings suggest that spleen-targeted ultrasound can blunt release of circulating cytokines and chemokines in the context of a human inflammatory disorder, without disrupting the overall immune cell repertoire.

### B Cells Retain Antibody Production Abilities

One concern in treating acute hyperinflammation is that the therapy may suppress the adaptive immune response, and thereby impair host protective immunity. Antibody production is a key protective function of B lymphocytes. Therefore, we assessed whether non-invasive splenic ultrasound treatment suppresses antibody production in RA patients. We observed that among genes upregulated after ultrasound treatment in RA patient B cells, are transcripts that encode for IgA, IgG, and IgM antibodies, under the GO term "B cell activation. Moreover, there is an enrichment for GO terms associated with “B cell activation” (Fig. 6A). We noted greater inter-subject variability in B cell gene expression changes (Fig. 6B), than we observed for suppression of cytokines and chemokines in monocytes. For example, we observed that for average transcript levels across B cells, transcripts encoding IgG are only upregulated in 3 subjects (IGHG2 and IGHG4, respectively) out of the 5 subjects. Regardless of the variability across subjects, these data support that ultrasound treatment does not generally inhibit B cell activation and antibody production across patients, and may even potentially enhance antibody production; thus, ultrasound is not expected to compromise humoral responses after short-term splenic ultrasound treatment.

**Figure 6.**
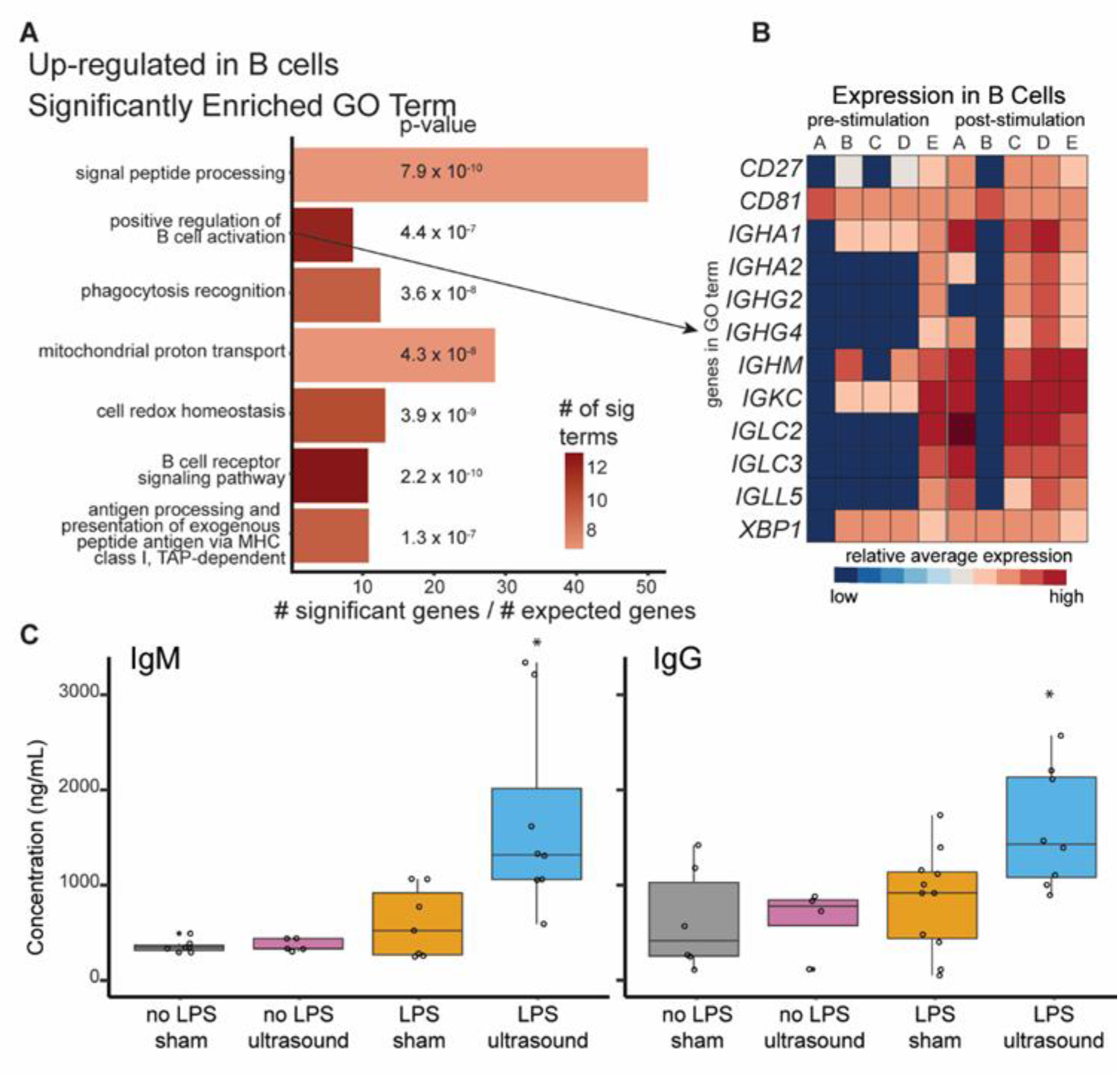
Ultrasound treatment does not reduce adaptive immune response. A) Enriched GO terms within upregulated genes in all B cells are shown, where bars represent number of significant genes in the list relative to number of genes that would be expected by chance, and with the number of genes contributing to each term represented by red hue. P-value for each term shown is based on Fisher exact test. B) Heatmap displaying genes that contribute to the GO term “positive regulation of B cell activation” within B cells. Each column represents relative average expression per subject for pre-stimulation (left 5 columns) and post-stimulation (right 5 columns) for each gene (labeled to the right). C) A rodent model of endotoxemia was used to further characterize the effects of splenic ultrasound stimulation on antibody production (further details of experiments are provided in Online Methods section). Box plot showing median levels of IgM (left) or IgG (right) concentrations in serum (open circles represent individual animals) for naïve animals (no LPS + sham), animals exposed to splenic ultrasound stimulation with no endotoxin (no LPS + ultrasound), animals exposed to the endotoxin but receiving sham stimulation (LPS + sham), and animals receiving both LPS exposure and splenic ultrasound stimulation (LPS + ultrasound). Animals receiving LPS + ultrasound showed significant increases in antibody production when compared to each of the other groups using an unpaired Wilcoxon ranked-sum test for IgM (no LPS + sham, p = 0.00145; no LPS + ultrasound, p = 0.00155; LPS + sham, p =0.01757) and for IgG (no LPS + sham, p =0.02930; no LPS + ultrasound, p = 0.00404; LPS + sham, p =0.04298). No other comparisons using this test showed a significance of p < 0.05.

To further investigate the effects of splenic ultrasound stimulation on adaptive immune response and antibody production we performed additional experiments using prophylactic ultrasound stimulation (i.e., ultrasound stimulation prior to antigen exposure) in a rodent model of endotoxemia (*29*). We observed that splenic ultrasound stimulation prior to endotoxin exposure (three minutes daily, once a day for 3 days followed by LPS injection on the fourth day) results in a significant ultrasound-induced increase in IgM and IgG antibody production 24 hours after LPS exposure compared to sham and/or non-LPS controls (p-values listed in Fig. 6C description). Furthermore, this enhanced antibody output is dependent on the presence of the toxin/antigen, as there is no observed ultrasound effect on antibody production in the absence of LPS exposure. This observation was further supported by a recent report demonstrating that brain-spleen neural pathways are capable of modulating splenic plasma cell differentiation upon presentation of novel T cell dependent antigens (*42*). Thus, providing a potential explanation for the bimodal distribution of human subjects that demonstrated an ultrasound-induced increase in antibody transcript levels, due to the neural pathway functional dependence on an active/on-going immune response.

## Discussion

By presenting first in-human data from two independent studies using different devices and protocols, we have demonstrated that non-invasive ultrasound stimulation of the spleen consistently drives anti-inflammatory effects in the context of an acute *ex vivo* response in healthy subjects and a chronic inflammatory condition. In RA patients after two weeks of daily 30-minute ultrasound treatment to the spleen, there was a reduction in cytokine IL-1β and chemokine IL-8 gene expression in circulating monocytes (Fig. 2). In healthy subjects, we observed a significant reduction in TNF protein levels produced by *ex vivo* LPS-stimulated whole blood, in individuals stimulated with 3 minutes of spleen ultrasound (Fig. 1B).

In addition to the reduction of key cytokines, we observed a reduction in activation of multiple signaling pathways in circulating monocytes with ultrasound treatment. Cytokines signal by binding to cell surface receptors to initiate a cascade of intracellular events. Important cytokines in the context of RA include TNF, IL-6, and IL-1β; their downstream intracellular pathways and necessary signaling molecules include cytokine receptors, components of the JAK/STAT pathway, MAP kinases and NFκB (*43, 44*). Many of the transcripts that encode for these critical components are downregulated with ultrasound treatment in RA patients (Fig. 2B). This finding suggests that in addition to cytokines and their immediate receptors, there is broad suppression of cellular pro-inflammatory pathways.

Down regulation of these various pro-inflammatory transcripts in RA patients occurs in the context of chronic inflammation. However, these same pathways are also involved in acute inflammatory responses (*44*). Genes induced by LPS-stimulated human monocytes include TNF, IL-1, IL-6, and IL-8 (*45*). NFκB and associated pathways are also upregulated, and NFκB is activated via phosphorylation during LPS exposure, allowing for a rapid induction of genes so the system can respond in an acute inflammatory setting. These same cellular components are downregulated with ultrasound treatment in our RA patients, suggesting that non-invasive splenic ultrasound could be used clinically to suppress acute inflammation as well.

The discovery that splenic ultrasound can reduce or blunt innate immune response in humans may have implications for patients suffering effects of overactive inflammatory response to infection and is quite timely in relation to the recent SARS-CoV-2 pandemic. Many of the same key inflammatory cytokines that are reduced with ultrasound stimulation are elevated in COVID- 19 patients. Recent studies have revealed that COVID-19 patients consistently overexpress IL-1β, IL-8, IL-6, and TNF, which strongly suggests dysregulation in the innate immune response (*46-49*). More specifically, SARS-CoV-2 infected individuals exhibit significantly higher expression levels of IL-1β and IL-6 transcripts in circulating monocytes, as well as IFNγ in CD8+ T cells, shown through PBMC single cell RNA sequencing of COVID-19 patients (*50*). These are the same pro-inflammatory molecules within the same cell types in which we observe an ultrasound-induced attenuation in gene expression. Consistent with this, single cell RNA sequencing has further shown that bronchoalveolar lavage fluid from patients with severe COVID-19 disease have higher levels of IL-8, IL-6 and IL-1β compared to patients with moderate disease (*51*). SARS-CoV-2 infection in children has been associated with a novel multisystem inflammatory disease (MIS-C) resembling toxic shock syndrome or atypical Kawasaki disease (*52*). MIS-C presents with elevated transcript levels of genes encoding for IL- 1, IL-6, IL-8, and IL-17, a condition caused by the COVID-19-induced cytokine storm (*53*). These molecules are key components of the COVID-19 cytokine storm that induce lung tissue damage and disease progression (*54*). Clinical therapies that can suppress these pro-inflammatory cytokines and reduce immune cells migration to the lungs could prove therapeutic for patients with moderate to severe COVID-19 disease, calming the cytokine storm and prevent morbidity and mortality (*55*) (Fig. S3). Additionally, recent reports suggest that targeting and reducing *multiple* factors involved in hyperinflammation (e.g. IL-6, IL-1β, IL-8, TNF, etc.) is a preferred strategy when attempting to dampen the hyperactive immune response, and combat COVID-19 (*56*). Here we show a novel therapy that can dampen several of these cytokines and downstream cellular pathways, using one form of noninvasive treatment (rather than multi-drug regimens).

In addition to demonstrating the ability to reduce pro-inflammatory cytokines and signaling pathways involved with a hyperactive innate immune response, splenic ultrasound reduces IL-8 and many components associated with cell migration, specifically monocyte migration (Fig. 4). These findings are consistent with previous animal studies showing that activation of the cholinergic anti-inflammatory pathway suppresses monocyte migration in mice (*43*). Taken together, one proposed mechanism of action for ultrasound treatment is that ultrasound modulates circulating immune cells initially within the spleen via activation of the cholinergic anti-inflammatory pathway, which suppresses pro-inflammatory cytokines and signaling pathways, as well as inhibiting cellular migration, such that there is reduced inflammation at the target site (e.g., at joints in RA or alveoli for COVID-19). Indeed such complimentary cholinergic anti-inflammatory pathway activities may contribute to the sustained disease modifying actions observed in RA and IBD clinical trials utilizing implant-based vagus nerve stimulators.

In addition to effects on innate immune function, data in humans and pre-clinical animal results demonstrate an immune state dependent increase in antibody production with splenic ultrasound. These results demonstrate an advantageous feature of splenic ultrasound compared to pharmaceutical options for immunosuppression, such as TNF, IL-6, or IL-1 blocking biologics (*57, 55*), in that cytokine suppression does not inhibit this other function of the adaptive immune system (i.e., B cell activity or antibody production). The potential pathway through which splenic ultrasound may boost the adaptive immune system response has recently been described, and features a brain-spleen neuroimmune pathway that enhances differentiation of antibody-producing splenic plasma cells (SPPCs) upon antigen exposure and activation of a novel immune system response (*42*). Therefore, splenic ultrasound stimulation may offer a new lower-risk and non-invasive approach for modulating the multiple factors in the innate and adaptive immune response, and provides a new tool to potentially treat a range of health conditions involving hyperinflammation, including infections and autoimmune diseases.

## Online Methods

### Splenic ultrasound in healthy subjects: Design of clinical study

For the splenic ultrasound study involving healthy human subjects (clinicaltrials.gov: NCT03548116), we provide the data from two cohorts: a sham control cohort (i.e., no ultrasound, with only transducer placement over the spleen) and a splenic ultrasound stimulation cohort (i.e., 290.4 mW/cm^2^ Ispta or 1.4 mechanical index (MI) using the shear wave elastography setting on the GE LOGIQ E9) that insonified a single site (i.e., the splenic hilum; as identified by a trained ultrasonographer performing the stimulation). Similar ultrasound parameters and targeting of the splenic hilum will be implemented for COVID-19 treatment in an upcoming clinical trial performed at the University of Minnesota together with GE Research funded by the Defense Advanced Research Projects Agency (DARPA; Department of Defense).

The testing of healthy subjects was carried out in accordance with the International Conference on Harmonisation Good Clinical Practice (ICH GCP) and the United States Code of Federal Regulations (CFR) applicable to clinical studies (45 CFR Part 46, 21 CFR Part 50, 21 CFR Part 56, and 21 CFR Part 812). The protocol, informed consent form, recruitment materials, and all participant materials were submitted and approved by the Institutional Review Board (IRB) at Northwell Health.

### Inclusion and exclusion criteria

To be eligible to participate in the study, an individual must have met the following criteria: aged between 18 and 45 years, without physical disability or conditions that may make them incapable of undergoing the study procedures or otherwise place them at greater harm, without significant past medical or surgical histories that would render them at a greater risk of harm, considered English proficient due to the study requirement to follow verbal commands during the ultrasound session, considered active as assessed by type of activity (e.g., walking or running) and number of hours a week performing the various activities, able to achieve all study requirements, able to comprehend the study goals and procedures, and able to provide informed consent for participation. See Table S5 for subject characteristics used in the cytokine analysis shown in Fig. 1.

### Study timeline

Subjects underwent a screening visit 1 to 11 days prior to baseline visit to assess eligibility to participate in the study. Eligible individuals that agreed to participate and provided written informed consent then underwent a physical and neurological examination. Women of childbearing potential were asked to provide a urine sample for pregnancy testing, and approximately 21 mL of blood was drawn. On the first protocol visit (day 0), participants again underwent a physical and neurological examination, and medical history review. A baseline blood draw was then collected for cytokine and blood chemistry analysis (~35 mL). The blood draw was performed under sterile conditions using standard venipuncture techniques. Individuals then underwent the ultrasound procedure based on random assignment to one of seven groups, including the two groups reported herein (i.e., the sham control group, and a stimulated group that received 100% power stimulus applied at the splenic target location). For ultrasound stimulus delivery, individuals were asked to lie in the right lateral recumbent position with their arms above their heads to expose the splenic region of the abdomen. After ultrasound gel was applied to the region, the ultrasound probe was placed on the participant’s abdomen. This procedure was performed for both the sham control group (n=9) and ultrasound stimulated group (n=9). Note that one subject from each group was excluded, in which 10 subjects were initially recruited to the study for each group (see Table S5).

### Stimulation paradigm

In the ultrasound stimulated cohort, the subject had the dimensions of their spleen assessed (using the B mode imaging setting on the GE LOGIQ E9), and the probe (C1-6) was positioned over the splenic hilum. Ultrasound application was performed using a modified elastography setting on the LOGIQ E9 operated in two modes: an imaging mode with short ultrasound pulses (on the order of 1 microsecond) and relatively low acoustic power levels (on the order of 10 mW/cm^2^) to initially target the spleen, and a stimulation mode (from the clinical shear wave elastography setting) using longer ultrasound pulses (on the order of 1 ms) and higher acoustic power levels (290.4 mW/cm^2^). Stimulation was performed in twelve 15- second long epochs separated by 15 seconds each, and a total stimulation duration of 3 minutes. The 15 second epochs were performed during breath holds, during which an ultrasonographer was holding the probe in position above the spleen. Before each stimulation period, the ultrasonographer checked the location of the probe using the imaging mode to ensure that all stimulation pulses were on target. The focal landmark used for targeting and stimulation was the hilum of the spleen. For the sham control group, the stimulation protocol and total duration of the procedure remained the same as the stimulated group, except that no ultrasound power was applied (i.e., ultrasound output via the probe was turned off). Two hours after the ultrasound stimulation protocol, another blood draw was performed.

### Blood analysis procedures

Prior to the *ex vivo* cytokine test, the LPS was sonicated for 30 minutes. A 10 mL blood tube (heparin; green cap) was brought into a sterile tissue culture room and blood was transferred to a 50 mL conical tube. A blinded researcher then diluted the LPS to the test concentrations in 15 pre-labeled tubes containing replicates of the different concentrations of LPS used for blood exposure, in which dilutions were made to achieve 0, 0.1, 1, and 10 ng/mL of LPS when mixed with the blood. Upon mixing the LPS with blood, the final tubes were capped and placed on a rocker within an incubator and the samples were kept at 37 degrees Celsius for 4 hours during incubation. After incubation, the samples were removed from the incubator and centrifuged at 6000 RPM for 5 minutes, and the plasma supernatant was transferred via pipette for storage in a -20°C freezer until analysis was performed. TNF analysis was performed in triplicate from each sample using a DuoSet ELISA kit (R&D Systems), as per manufacturer’s instructions.

### Splenic ultrasound in RA patients: Design of clinical study

The study is a double-blinded, controlled and randomized clinical trial investigating the therapeutic effects of a 14-day regimen of splenic ultrasound stimulation in RA patients (listed at clinicaltrials.gov (NCT03690466) for which a portion of the results are presented in this paper. The clinical trial is still ongoing, in which the primary and secondary outcome measures evaluating within-arm and between-arm DAS-28-CRP comparisons for the treated and sham groups will be reported in a subsequent paper after the completion of the study. Due to the urgent need for identifying new treatments for SARS-CoV-2 and to guide an upcoming clinical trial in COVID-19 patients at the University of Minnesota funded by the Defense Advanced Research Projects Agency (DARPA; Department of Defense), additional single cell RNA sequencing and data analysis were performed for five ultrasound stimulated patients in the study. This analysis was motivated by recently published data on single cell RNA sequencing in COVID-19 patients showing which cytokines in PBMCs are associated with their life-threatening symptoms (*50*), and has revealed that splenic ultrasound stimulation can suppress pro-inflammatory pathways relevant for treating SARS-CoV-2 infection and other hyperinflammation conditions.

The protocol, informed consent form, recruitment materials, and all participant materials were submitted and approved by the University of Minnesota’s Institutional Review Board (IRB) and monitored by CTSI in accordance with its institutionally approved monitoring plan. The description below includes relevant methods and analyses for the results presented in this paper.

### Inclusion and exclusion criteria

To be eligible to participate in the study, an individual must have met the following criteria: 18 years of age or older, a diagnosis of seropositive RA (rheumatoid factor-positive or cyclic citrullinated peptide antibody-positive), and symptoms or signs of inadequate disease control (either modified HAQ score >0.3 or DAS-28-CRP >3.2). Exclusion criteria included active bacterial or viral infection, pregnancy, malignancy, or inability to provide daily self-care. The patient medical history and characteristics for each of the five subjects presented in this paper are provided in Table S2.

### Study timeline

Participants were assessed during in-clinic visits on days 0 (before treatment began), 3, 7, 10, 14, and 21. Minor flexibility was allowed for scheduling clinic visits outside of holidays for special circumstances. On the first visit, the spleen was imaged by an ultrasonographer and the center of the spleen trajectory was marked on the participant’s skin. Participants were instructed on how to self-administer ultrasound treatment to the spleen. The procedure involved sweeping the ultrasound wand continuously across a 5-inch by 5-inch square centered on the spleen mark placed by the ultrasonographer and ensuring full contact was made with the skin during the daily 30 minute stimulation period. Researchers were able to visually confirm that daily treatments were properly administered through daily online video sessions with the patients. These video sessions took place at about the same time each day for the 14-day treatment period. With each in-clinic visit, participants received a joint examination, peripheral joint ultrasound, and blood draw, as well as completing several physical evaluations and questionnaires. Results for all patients and assessments will be presented in a future publication after recruitment of 20 total subjects and completion of the study.

### Treatment

After an in-clinic training session on Day 0, transcutaneous ultrasound was administered to the spleen for 30 minutes daily for 2 weeks (14 days total) via a portable device (1.2 W/cm^2^; SoundCare Plus, Roscoe Medical) in the patient’s home. The patients are randomized (1:1) to a treatment group or a control (sham) group, in which the latter receives the exact same evaluations and stimulation paradigm as the treatment group, except that the device does not deliver any energy from the ultrasound transducer. RA patients and clinical assessors are blinded as to which patients received ultrasound or sham treatment.

### RNA sequencing and statistical analyses

4mL of blood was collected in a green top LiHep tube from each ultrasound stimulated participant (n=5) on day 0 (before the first splenic ultrasound stimulation was administered) and on day 14 (after the final treatment session). The samples were processed the same day as collection. Peripheral blood mononuclear cells were isolated from whole blood using density gradient centrifugation via SepMate tubes (Stemcell Technologies) and following manufacturer’s instructions. PBMCs were then gently frozen and stored at -80 °C as per 10X Genomics demonstrated protocols GC000039. Following this protocol, once all samples had been collected and frozen, samples were thawed quickly in 20% FBS in PBS, resuspended in 10% FBS, and strained before proceeding to 10X genomics single cell protocol. Samples were sequenced using UMI-based approach from10X Genomics, as previously described (*30*) and mapped to the human genome.

Transcriptomic analysis was performed using R statistical software, and primary filtering and normalization of each sample was done using Seurat package as previously described (*30, 59, 60*). Samples included two timepoints from five subjects and these individual samples were merged for cell type assignment and differential expression analysis. Cell types were assigned using standard marker genes (see Fig. S2) as previously described (*61*). Differential expression was determined between timepoints for all cells in each cell type using a negative binomial generalized linear model. GO term analysis was performed using the org.Hs.eg.db and topGO packages. GO term enrichment was assessed using the ‘elim’ algorithm with a Fisher exact test cutoff of 0.01, in which the top 40 significant GO terms in monocytes are shown in Table S3 (*62*).

### Pre-clinical tests in LPS/endotoxin rodent model

Pre-clinical tests in rodents were performed as previously described (*29*). Experiments were performed under protocols approved by the Institutional Animal Care and Use Committee of GE Research. Briefly, a VIVID E9 (GE) ultrasound system was used to perform an ultrasound scan and locate the spleen. A HIFU transducer and system was then positioned on the target area, and a separate ultrasound scan performed using a smaller probe (3S, GE) that was placed within the opening of the HIFU transducer was used to verify alignment of the ultrasound beam with the spleen. Adult Sprague-Dawley rats that were 8-12 weeks old (250-300 g; Charles River Labs) were housed at 25 degrees Celsius on a 12-h light/dark cycle and acclimatized for 1 week, with handling before experiments to minimize the potential confounding measures due to stress response. Water and regular rodent chow were available ad libitum. LPS (0111:B4, Sigma Aldrich) was used to produce a significant state of inflammation in the naïve adult rodents. LPS was administered to animals in the amount of 10 mg/kg, which corresponds to a LD75 dose via intraperitoneal injection.

### Treatment

For ultrasound stimulation, animals were anesthetized with 2-4% isoflurane, and the animals were laid on a water circulating warming pad to prevent hyperthermia during the procedures. The region designated for ultrasound stimulation was shaved with a disposable razor and animal clippers prior to stimulation. Ultrasound was applied to the designated area above the spleen using the probe and system described and calibrated from a previous study (*29*). An ultrasound stimulus using the previously found optimal stimulation parameters (1.1 MHz, 0.5 ms pulse repetition frequency, 136.36 microsecond pulse length, 568 mW/cm^2^ Ispta) was then applied for 2 minutes, and LPS was immediately injected post-stimulation. Animals were allowed to incubate under anesthesia for one hour. After incubation, the animal was euthanized and tissue and blood samples were collected. For experiments testing antibody production, an ultrasound stimulus was applied daily (in the morning) for three days prior to LPS injection. On the fourth day, following the three consecutive treatment days, LPS (10mg/kg) was injected intraperitoneally. The animals were then incubated for 60 minutes in accordance with previous studies (*29*) and tissue and blood were harvested for analysis of cytokines and antibody production.

### Tissue collection

An incision was made starting at the base of the peritoneal cavity extending up and through to the pleural cavity. The spleen was rapidly removed and homogenized in a solution of phosphate-buffered saline, containing phosphatase (0.2-mM phenylmethylsulfonyl fluoride, 5-μg/mL aprotinin, 1-mM benzamidine, 1-mM sodium orthovanadate, and 2-μM cantharidin) and protease (1-μL to 20 mg of tissue as per Roche Diagnostics) inhibitors. A targeted final concentration of 0.2-g tissue per mL PBS solution was applied in all samples. After collection of the whole blood, we allowed the blood to clot undisturbed at room temperature for 15-30 minutes. Samples were centrifuged at 1,000-2,000x g for 10 minutes in a refrigerated centrifuge with the resulting supernatant being collected for antibody testing. Samples were then stored at -80 °C until analysis. Splenic tissue lysate was analyzed for cytokine concentrations using ELISA kits for quantifying TNF alpha (Abcam, Ab100785), IFN gamma (Abcam, Ab239425), IL-13 (Abcam, Ab100766), IL-12 (LSBio, LS-F34357), IL-10 (Abcam, Ab100765), IL-2 (Abcam, Ab221834), IL-1 beta (Abcam, Ab100768), IL-1 alpha (Abcam, Ab113350), IL-6 (Abcam, Ab234570) and IL-4 (Abcam, Ab100771) as per manufacturer’s instructions for tissue samples. Serum was tested using ELISA kits for quantifying antibody production including IgG (Abcam, Ab189578), and IgM (Abcam, Ab215085) as per manufacturer’s instructions.

## Data Availability

Data associated with this study are presented in this paper or supplemental document. The RNA-seq data presented in this paper will soon be deposited in the Gene Expression Omnibus database. Other data or methods such as that used for bioinformatic analyses or visualization are available upon reasonable request. 

## Acknowledgments

We thank all study participants. We are grateful for the assistance of individuals at the University of Minnesota Genomics Center, especially Jerry Daniel and Emma Stanley. Additionally, we would like to thank Stuart Sealfon and Frederique Ruf-zamojski for their assistance and advice processing PBMC samples for single cell RNA sequencing. This work was supported by the United States Defense Advanced Research Projects Agency (DARPA) Electrical Prescriptions (ElectRx) Program under the guidance of Eric Van Gieson and Gretchen Knaack at DARPA. The views, opinions and/or findings expressed are those of the authors and should not be interpreted as representing the official views or policies of the Department of Defense or the U.S. Government.

## Author Contributions

For research associated with splenic ultrasound in healthy participants and in rodents: K.W., J.A., C.P., R.R., S.S.C., T.R.C., J.P., K.J.T., S.Z. contributed to study design and setup; V.C., J.G., C.D’A., D.N., J.P. performed data collection and analysis; and C.P., V.C., T.R.C. contributed to data interpretation and writing. For research associated with splenic ultrasound of rheumatoid arthritis patients: R.S.G., D.P.Z., C.R.W.K., B.A.B., E.P., H.H.L. contributed to study design and setup; R.S.G., D.P.Z., C.R.W.K., E.P. performed data collection and analysis; R.S.G. performed data collection and bioinformatic analysis of single cell transcriptomic data; and R.S.G., D.P.Z., B.A.B., E.P., H.H.L contributed to data interpretation and writing. R.S.G., H.H.L. contributed to the first draft of the manuscript. All authors contributed to and approved the final version of the manuscript.

## Data and Materials availability

Data associated with this study are presented in this paper or supplemental document. The single cell RNA sequencing data presented in this paper have been deposited in the Gene Expression Omnibus database as GSExxxxxx (currently being submitted). Other data or methods such as that used for bioinformatic analyses or visualization are available upon reasonable request.

## Supplementary

**Figure S1.**
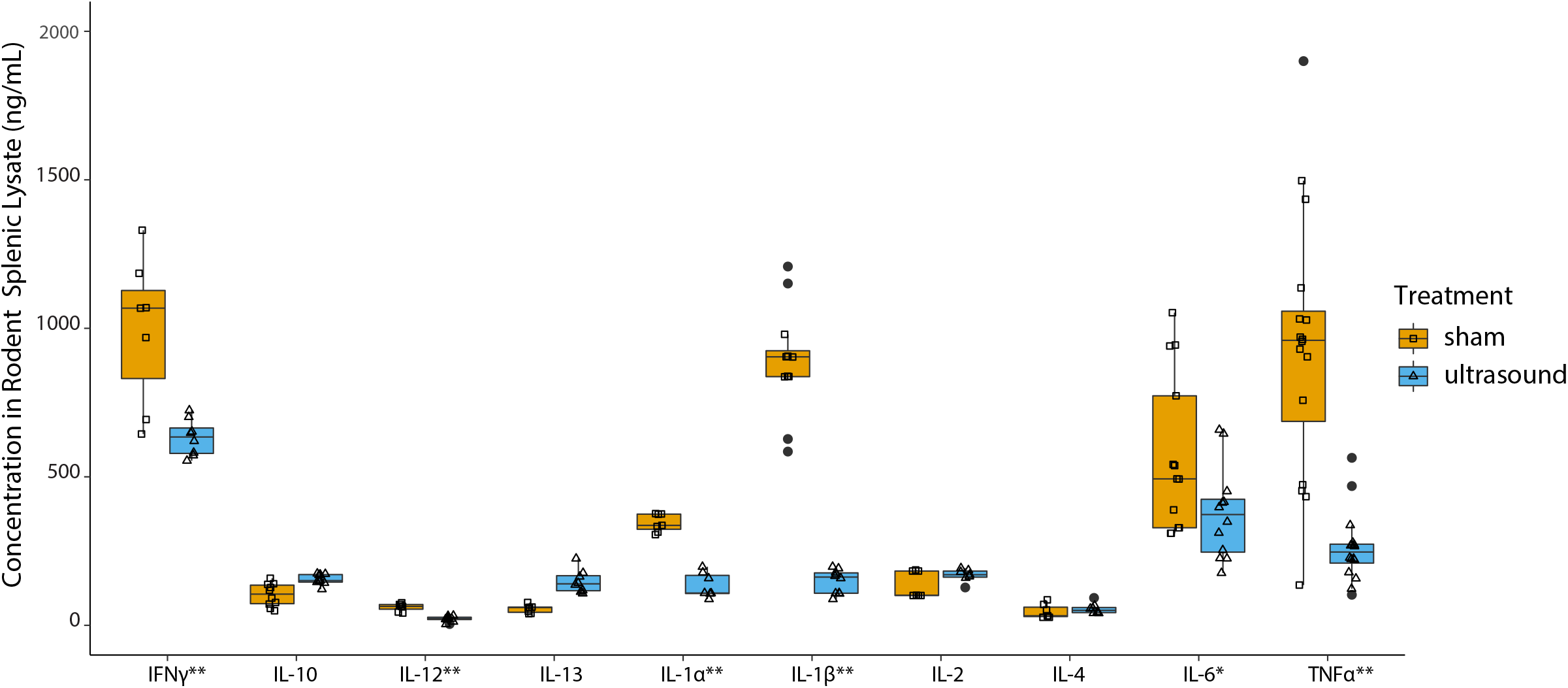
Modulation of Cytokines with Splenic Ultrasound in Rats. Splenic ultrasound was applied to rats for 2 minutes daily for 3 days prior to the day of intraperitoneal LPS injection. Boxplot shows cytokine levels in splenic lysates from animals 1 hour post LPS injection receiving ultrasound or sham stimulation. Sham treated animals are shown with orange boxes with open squares representing individual data points, ultrasound stimulated animals are shown with blue boxes with open triangles representing individual datapoints and black dots represent outliers. Cytokines that show a significant reduction with ultrasound are labeled with one asterisk based on a one-sided unpaired Wilcoxon rank-sum test (p<0.05; expecting a reduction in cytokines). Cytokines with two asterisks correspond to a p<0.005. Significant reduction is observed in IFNγ, IL-12, IL-1α, IL-1β, IL-6, and TNF.

| Cytokine | # sham samples | # stim samples | one-sided unpaired Wilcoxon rank sum p-value |
| --- | --- | --- | --- |
| IFNγ | 7 | 8 | 0.004662 |
| IL-10 | 10 | 9 | 0.999500 |
| IL-12 | 7 | 9 | 0.000508 |
| IL-13 | 7 | 8 | 1.000000 |
| IL-1α | 7 | 7 | 0.000291 |
| IL-1β | 11 | 9 | 0.000002 |
| IL-2 | 7 | 7 | 0.875200 |
| IL-4 | 7 | 8 | 0.884100 |
| IL-6 | 12 | 12 | 0.034160 |
| TNF | 18 | 16 | 0.000001 |

**Figure S2.**
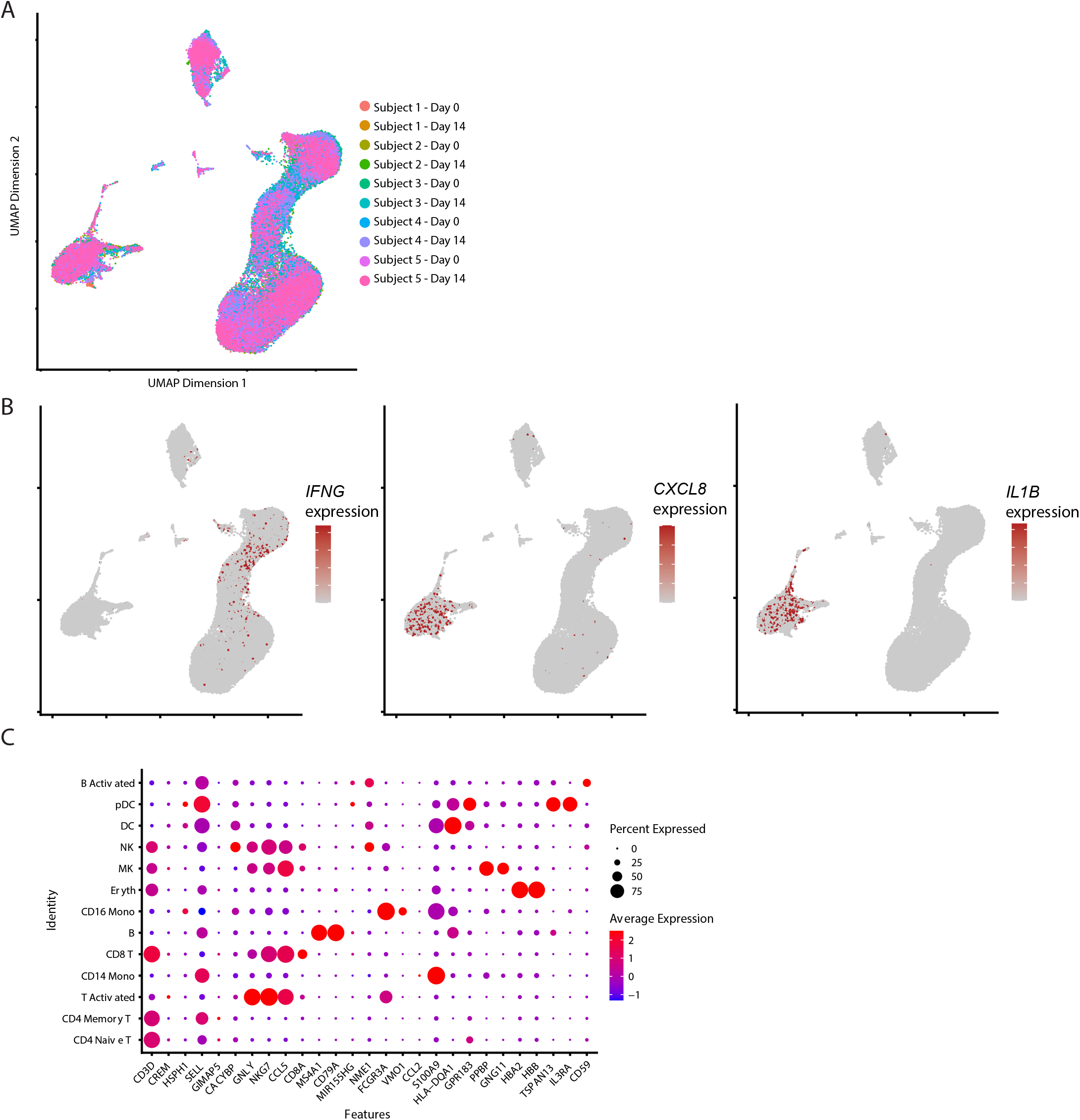
Cellular Transcriptional Signatures. Single cell RNA sequencing results on peripheral blood mononuclear cells. Uniform Manifold Approximation and Projection (UMAP) dimensionality reduction was used on principal component analysis that determines which gene sets contribute the most variability among the dataset. A. UMAP plot of cells from each subject and timepoint to show transcriptional similarities between subjects and timepoints. B. UMAP plot shows cells expressing IFNG, CXCL8 and IL1B transcripts, and their expression patterns within particular cell types match with which cell types show statistical differential expression for those transcripts. C. Dotplot showing the expression (color) and percent of cells expressing (dot size) each gene transcript (x-axis) for each cell type (y-axis). The genes listed on the x-axis are commonly used marker genes that help determine and assign cell types to groups of cells from an unbiased principal component analysis.

**Figure S3.**
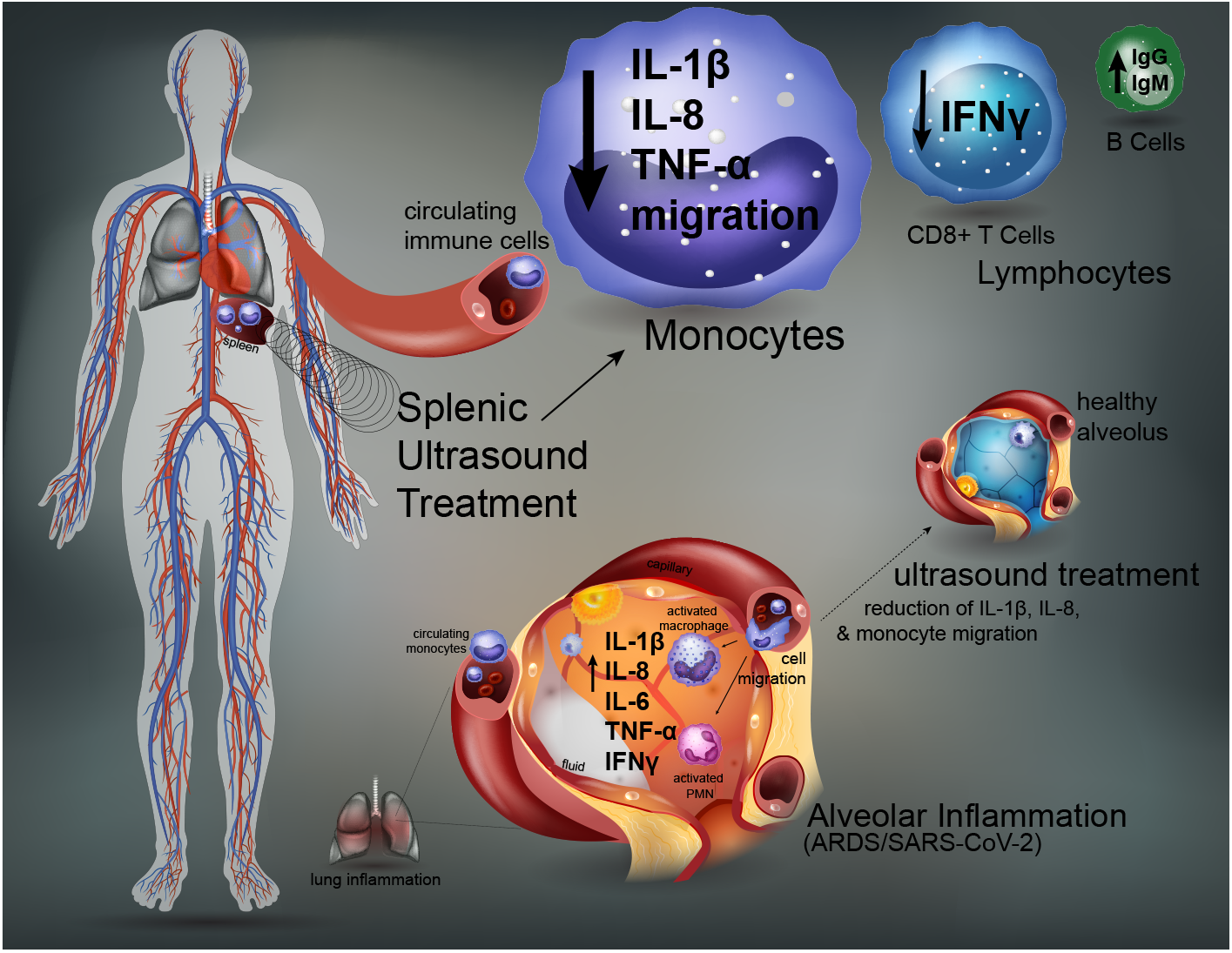
Non-invasive splenic ultrasound treatment for ameliorating immune hyperactivity for COVID-19. Schematic of the mechanism of splenic ultrasound treatment's effect on circulating immune cells. Splenic ultrasound reduces IL-1β, IL-8, TNF and genes involved in monocyte migration in monocytes, reduces IFNγ in CD8+ T cells, and increased IgG and IgM production in B cells (upper left, solid arrow). This treatment approach could help combat key elevated cytokines in alveoli and/or cellular migration to lungs in the context of COVID-19 (lower right, dashed arrow).

**Table S1.** Single Cell RNA sequencing: Number of Cells, Reads, and Genes. Table showing the number of cells for each cell type, which were determined by gene signatures (see Fig. S2C), and also showing number of reads (counts) and average number of genes for each cell type. We observe that all cells have a relatively consistent read depth for the sequencing reaction performed.

| celltype | # of cells | total counts | average counts/cell | average genes/cell |
| --- | --- | --- | --- | --- |
| B Activated day 0 | 20 | 1.57E+05 | 7.85E+03 | 2171 |
| B Activated day 14 | 24 | 1.53E+05 | 6.38E+03 | 1815 |
| B day 0 | 2485 | 8.87E+06 | 3.57E+03 | 1134 |
| B day 14 | 1835 | 6.88E+06 | 3.75E+03 | 1151 |
| CD14 Mono day 0 | 3465 | 1.69E+07 | 4.87E+03 | 1878 |
| CD14 Mono day 14 | 2150 | 9.77E+06 | 4.54E+03 | 1743 |
| CD16 Mono day 0 | 434 | 2.77E+06 | 6.37E+03 | 2184 |
| CD16 Mono day 14 | 324 | 1.97E+06 | 6.08E+03 | 2073 |
| CD4 Memory T day 0 | 8568 | 2.60E+07 | 3.03E+03 | 955 |
| CD4 Memory T day 14 | 4063 | 1.16E+07 | 2.86E+03 | 914 |
| CD4 Naive T day 0 | 7206 | 2.55E+07 | 3.55E+03 | 1154 |
| CD4 Naive T day 14 | 3629 | 1.17E+07 | 3.21E+03 | 1080 |
| CD8 T day 0 | 5764 | 1.63E+07 | 2.83E+03 | 1095 |
| CD8 T day 14 | 2505 | 7.78E+06 | 3.11E+03 | 1178 |
| DC day 0 | 138 | 1.19E+06 | 8.62E+03 | 2403 |
| DC day 14 | 102 | 8.41E+05 | 8.24E+03 | 2365 |
| Eryth day 0 | 388 | 1.19E+06 | 3.06E+03 | 875 |
| Eryth day 14 | 356 | 1.34E+06 | 3.77E+03 | 993 |
| MK day 0 | 288 | 7.48E+05 | 2.60E+03 | 1027 |
| MK day 14 | 130 | 3.40E+05 | 2.62E+03 | 1063 |
| NK day 0 | 78 | 4.30E+05 | 5.51E+03 | 1956 |
| NK day 14 | 192 | 1.14E+06 | 5.93E+03 | 1991 |
| pDC day 0 | 49 | 2.85E+05 | 5.82E+03 | 2080 |
| pDC day 14 | 74 | 3.94E+05 | 5.32E+03 | 1921 |
| T Activated day 0 | 5545 | 1.46E+07 | 2.63E+03 | 1162 |
| T Activated day 14 | 3531 | 9.02E+06 | 2.55E+03 | 1144 |
| **Total** | **53343** | **1.78E+08** |  |  |

**Table S2.** Splenic Ultrasound in RA Patients: Subject Features. Table showing key information for each participant, including age, sex, race, and medications the participants continued taking during the study. For figures in this publication where gene expression averages are shown per participant, participants are also shown as columns corresponding to A to E in this table.

| Medical History and RA Features (at time of first visit) | Participant A | Participant B | Participant C | Participant D | Participant E |
| --- | --- | --- | --- | --- | --- |
| Visit 1 Date | 4/18/2019 | 5/23/2019 | 8/29/2019 | 12/2/2019 | 12/2/2019 |
| Age | 68 | 57 | 89 | 42 | 41 |
| Sex | Female | Female | Male | Female | Female |
| Ethnic Group (as indicated in medical records) | American | American | American | Hmong | "Patient chose not to answer" |
| Height (inches) | 60 | 65 | 67 | 56 | 65 |
| Weight (lbs) | 180 | 139 | 170 | 118 | 256 |
| Body Mass Index | 35.2 | 23.1 | 26.6 | 26.5 | 43.3 |
| Date of RA diagnosis | 3/4/2019 | 6/1/2014 | 8/20/2019 | 1/1/2013 | 12/6/2013 |
| Time since RA diagnosis | 1 month | 4 years | 1 week | 6 years | 5 years |
| Prescription medicine for RA immune modulation which participants continued taking during study | Prednisone | None | None | Hydroxychloroquine | Orencia, methotrexate |
| Prescription medicine for pain taken during study | None | Hydrocodone | None | None | None |
| Non-prescription pain medicine taken during study | Acetaminophen | None | Ibuprofen, Aspirin | Acetaminophen, Naproxen | Naproxen |

**Table S3.**
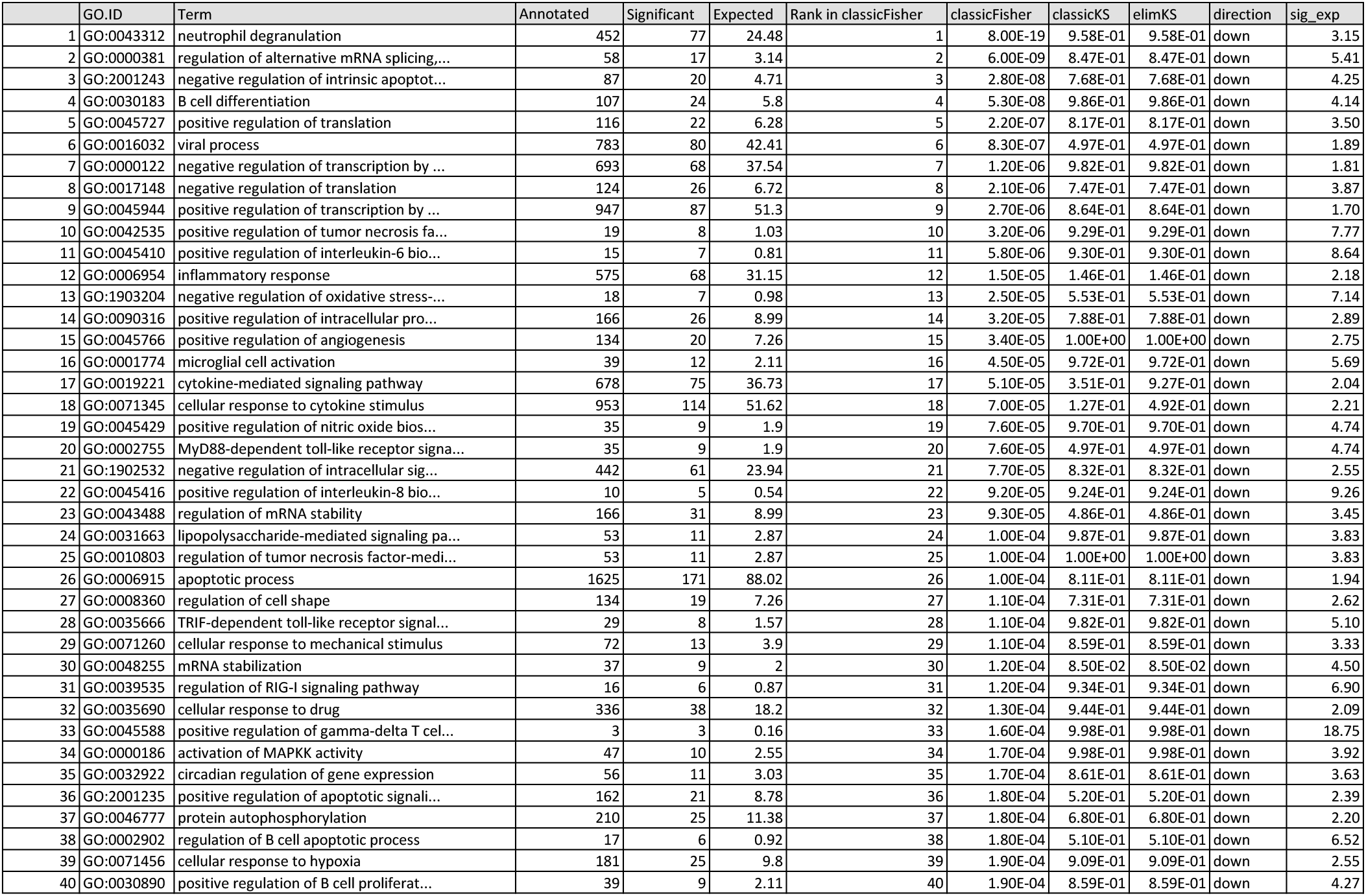
Top 40 Enriched GO Terms for Downregulated Genes in Monocytes. Table displays enriched GO terms for genes downregulated with ultrasound treatment. Also shown are the number of genes out of the human genome that are annotated with that GO term, number of genes from the list of significantly downregulated genes with ultrasound, and the number of genes that would be expected by random chance. Column labeled 'classicFisher' shows p-values from the Fisher's exact test and 'sig_exp' is the number of genes found to be significant in our gene list (decreased with ultrasound in monocytes) compared to the number that might be expected by chance.

|  | GO.ID | Term | Annotated | Significant | Expected | Rank in classicFisher | classicFisher | classicKS | elimKS | direction | sig_exp |
| --- | --- | --- | --- | --- | --- | --- | --- | --- | --- | --- | --- |
| 1 | GO:0043312 | neutrophil degranulation | 452 | 77 | 24.48 | 1 | 8.00E-19 | 9.58E-01 | 9.58E-01 | down | 3.15 |
| 2 | GO:0000381 | regulation of alternative mRNA splicing,... | 58 | 17 | 3.14 | 2 | 6.00E-09 | 8.47E-01 | 8.47E-01 | down | 5.41 |
| 3 | GO:2001243 | negative regulation of intrinsic apoptot... | 87 | 20 | 4.71 | 3 | 2.80E-08 | 7.68E-01 | 7.68E-01 | down | 4.25 |
| 4 | GO:0030183 | B cell differentiation | 107 | 24 | 5.8 | 4 | 5.30E-08 | 9.86E-01 | 9.86E-01 | down | 4.14 |
| 5 | GO:0045727 | positive regulation of translation | 116 | 22 | 6.28 | 5 | 2.20E-07 | 8.17E-01 | 8.17E-01 | down | 3.50 |
| 6 | GO:0016032 | viral process | 783 | 80 | 42.41 | 6 | 8.30E-07 | 4.97E-01 | 4.97E-01 | down | 1.89 |
| 7 | GO:0000122 | negative regulation of transcription by ... | 693 | 68 | 37.54 | 7 | 1.20E-06 | 9.82E-01 | 9.82E-01 | down | 1.81 |
| 8 | GO:0017148 | negative regulation of translation | 124 | 26 | 6.72 | 8 | 2.10E-06 | 7.47E-01 | 7.47E-01 | down | 3.87 |
| 9 | GO:0045944 | positive regulation of transcription by ... | 947 | 87 | 51.3 | 9 | 2.70E-06 | 8.64E-01 | 8.64E-01 | down | 1.70 |
| 10 | GO:0042535 | positive regulation of tumor necrosis fa... | 19 | 8 | 1.03 | 10 | 3.20E-06 | 9.29E-01 | 9.29E-01 | down | 7.77 |
| 11 | GO:0045410 | positive regulation of interleukin-6 bio... | 15 | 7 | 0.81 | 11 | 5.80E-06 | 9.30E-01 | 9.30E-01 | down | 8.64 |
| 12 | GO:0006954 | inflammatory response | 575 | 68 | 31.15 | 12 | 1.50E-05 | 1.46E-01 | 1.46E-01 | down | 2.18 |
| 13 | GO:1903204 | negative regulation of oxidative stress-... | 18 | 7 | 0.98 | 13 | 2.50E-05 | 5.53E-01 | 5.53E-01 | down | 7.14 |
| 14 | GO:0090316 | positive regulation of intracellular pro... | 166 | 26 | 8.99 | 14 | 3.20E-05 | 7.88E-01 | 7.88E-01 | down | 2.89 |
| 15 | GO:0045766 | positive regulation of angiogenesis | 134 | 20 | 7.26 | 15 | 3.40E-05 | 1.00E+00 | 1.00E+00 | down | 2.75 |
| 16 | GO:0001774 | microglial cell activation | 39 | 12 | 2.11 | 16 | 4.50E-05 | 9.72E-01 | 9.72E-01 | down | 5.69 |
| 17 | GO:0019221 | cytokine-mediated signaling pathway | 678 | 75 | 36.73 | 17 | 5.10E-05 | 3.51E-01 | 9.27E-01 | down | 2.04 |
| 18 | GO:0071345 | cellular response to cytokine stimulus | 953 | 114 | 51.62 | 18 | 7.00E-05 | 1.27E-01 | 4.92E-01 | down | 2.21 |
| 19 | GO:0045429 | positive regulation of nitric oxide bios... | 35 | 9 | 1.9 | 19 | 7.60E-05 | 9.70E-01 | 9.70E-01 | down | 4.74 |
| 20 | GO:0002755 | MyD88-dependent toll-like receptor signa... | 35 | 9 | 1.9 | 20 | 7.60E-05 | 4.97E-01 | 4.97E-01 | down | 4.74 |
| 21 | GO:1902532 | negative regulation of intracellular sig... | 442 | 61 | 23.94 | 21 | 7.70E-05 | 8.32E-01 | 8.32E-01 | down | 2.55 |
| 22 | GO:0045416 | positive regulation of interleukin-8 bio... | 10 | 5 | 0.54 | 22 | 9.20E-05 | 9.24E-01 | 9.24E-01 | down | 9.26 |
| 23 | GO:0043488 | regulation of mRNA stability | 166 | 31 | 8.99 | 23 | 9.30E-05 | 4.86E-01 | 4.86E-01 | down | 3.45 |
| 24 | GO:0031663 | lipopolysaccharide-mediated signaling pa... | 53 | 11 | 2.87 | 24 | 1.00E-04 | 9.87E-01 | 9.87E-01 | down | 3.83 |
| 25 | GO:0010803 | regulation of tumor necrosis factor-medi... | 53 | 11 | 2.87 | 25 | 1.00E-04 | 1.00E+00 | 1.00E+00 | down | 3.83 |
| 26 | GO:0006915 | apoptotic process | 1625 | 171 | 88.02 | 26 | 1.00E-04 | 8.11E-01 | 8.11E-01 | down | 1.94 |
| 27 | GO:0008360 | regulation of cell shape | 134 | 19 | 7.26 | 27 | 1.10E-04 | 7.31E-01 | 7.31E-01 | down | 2.62 |
| 28 | GO:0035666 | TRIF-dependent toll-like receptor signal... | 29 | 8 | 1.57 | 28 | 1.10E-04 | 9.82E-01 | 9.82E-01 | down | 5.10 |
| 29 | GO:0071260 | cellular response to mechanical stimulus | 72 | 13 | 3.9 | 29 | 1.10E-04 | 8.59E-01 | 8.59E-01 | down | 3.33 |
| 30 | GO:0048255 | mRNA stabilization | 37 | 9 | 2 | 30 | 1.20E-04 | 8.50E-02 | 8.50E-02 | down | 4.50 |
| 31 | GO:0039535 | regulation of RIG-I signaling pathway | 16 | 6 | 0.87 | 31 | 1.20E-04 | 9.34E-01 | 9.34E-01 | down | 6.90 |
| 32 | GO:0035690 | cellular response to drug | 336 | 38 | 18.2 | 32 | 1.30E-04 | 9.44E-01 | 9.44E-01 | down | 2.09 |
| 33 | GO:0045588 | positive regulation of gamma-delta T cel... | 3 | 3 | 0.16 | 33 | 1.60E-04 | 9.98E-01 | 9.98E-01 | down | 18.75 |
| 34 | GO:0000186 | activation of MAPKK activity | 47 | 10 | 2.55 | 34 | 1.70E-04 | 9.98E-01 | 9.98E-01 | down | 3.92 |
| 35 | GO:0032922 | circadian regulation of gene expression | 56 | 11 | 3.03 | 35 | 1.70E-04 | 8.61E-01 | 8.61E-01 | down | 3.63 |
| 36 | GO:2001235 | positive regulation of apoptotic signali... | 162 | 21 | 8.78 | 36 | 1.80E-04 | 5.20E-01 | 5.20E-01 | down | 2.39 |
| 37 | GO:0046777 | protein autophosphorylation | 210 | 25 | 11.38 | 37 | 1.80E-04 | 6.80E-01 | 6.80E-01 | down | 2.20 |
| 38 | GO:0002902 | regulation of B cell apoptotic process | 17 | 6 | 0.92 | 38 | 1.80E-04 | 5.10E-01 | 5.10E-01 | down | 6.52 |
| 39 | GO:0071456 | cellular response to hypoxia | 181 | 25 | 9.8 | 39 | 1.90E-04 | 9.09E-01 | 9.09E-01 | down | 2.55 |
| 40 | GO:0030890 | positive regulation of B cell proliferat... | 39 | 9 | 2.11 | 40 | 1.90E-04 | 8.59E-01 | 8.59E-01 | down | 4.27 |

**Table S4.**
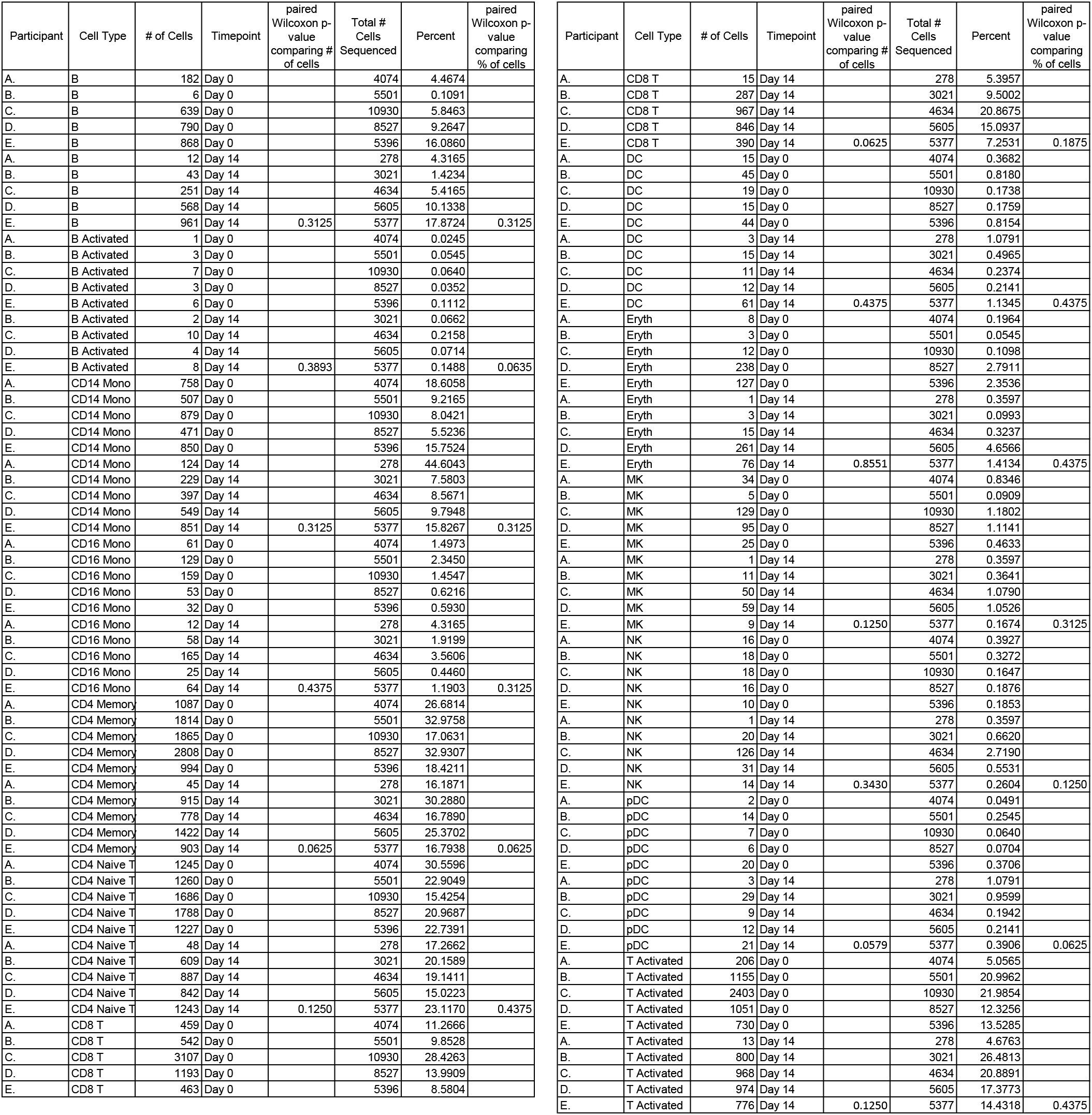
Cell Type Counts Per Participant and Statistics. Table displays the number of cells sequenced and percent (# cells in cell type / # cells sequenced in that sample x 100) for each sample (defined as a participant and timepoint) per cell type. No significant difference is seen in the number of cells sequenced per cell type or percent within each cell type comparing pre- and post ultrasound stimulation with a paired Wilcoxon rank sum test that was used to determine p-values.

| Participant | Cell Type | # of Cells | Timepoint | paired Wilcoxon p-value comparing # of cells | Total # Cells Sequenced | Percent | paired Wilcoxon p-value comparing % of cells |
| --- | --- | --- | --- | --- | --- | --- | --- |
| A. | B | 182 | Day 0 |  | 4074 | 4.4674 |  |
| B. | B | 6 | Day 0 |  | 5501 | 0.1091 |  |
| C. | B | 639 | Day 0 |  | 10930 | 5.8463 |  |
| D. | B | 790 | Day 0 |  | 8527 | 9.2647 |  |
| E. | B | 868 | Day 0 |  | 5396 | 16.0860 |  |
| A. | B | 12 | Day 14 |  | 278 | 4.3165 |  |
| B. | B | 43 | Day 14 |  | 3021 | 1.4234 |  |
| C. | B | 251 | Day 14 |  | 4634 | 5.4165 |  |
| D. | B | 568 | Day 14 |  | 5605 | 10.1338 |  |
| E. | B | 961 | Day 14 | 0.3125 | 5377 | 17.8724 | 0.3125 |
| A. | B Activated | 1 | Day 0 |  | 4074 | 0.0245 |  |
| B. | B Activated | 3 | Day 0 |  | 5501 | 0.0545 |  |
| C. | B Activated | 7 | Day 0 |  | 10930 | 0.0640 |  |
| D. | B Activated | 3 | Day 0 |  | 8527 | 0.0352 |  |
| E. | B Activated | 6 | Day 0 |  | 5396 | 0.1112 |  |
| B. | B Activated | 2 | Day 14 |  | 3021 | 0.0662 |  |
| C. | B Activated | 10 | Day 14 |  | 4634 | 0.2158 |  |
| D. | B Activated | 4 | Day 14 |  | 5605 | 0.0714 |  |
| E. | B Activated | 8 | Day 14 | 0.3893 | 5377 | 0.1488 | 0.0635 |
| A. | CD14 Mono | 758 | Day 0 |  | 4074 | 18.6058 |  |
| B. | CD14 Mono | 507 | Day 0 |  | 5501 | 9.2165 |  |
| C. | CD14 Mono | 879 | Day 0 |  | 10930 | 8.0421 |  |
| D. | CD14 Mono | 471 | Day 0 |  | 8527 | 5.5236 |  |
| E. | CD14 Mono | 850 | Day 0 |  | 5396 | 15.7524 |  |
| A. | CD14 Mono | 124 | Day 14 |  | 278 | 44.6043 |  |
| B. | CD14 Mono | 229 | Day 14 |  | 3021 | 7.5803 |  |
| C. | CD14 Mono | 397 | Day 14 |  | 4634 | 8.5671 |  |
| D. | CD14 Mono | 549 | Day 14 |  | 5605 | 9.7948 |  |
| E. | CD14 Mono | 851 | Day 14 | 0.3125 | 5377 | 15.8267 | 0.3125 |
| A. | CD16 Mono | 61 | Day 0 |  | 4074 | 1.4973 |  |
| B. | CD16 Mono | 129 | Day 0 |  | 5501 | 2.3450 |  |
| C. | CD16 Mono | 159 | Day 0 |  | 10930 | 1.4547 |  |
| D. | CD16 Mono | 53 | Day 0 |  | 8527 | 0.6216 |  |
| E. | CD16 Mono | 32 | Day 0 |  | 5396 | 0.5930 |  |
| A. | CD16 Mono | 12 | Day 14 |  | 278 | 4.3165 |  |
| B. | CD16 Mono | 58 | Day 14 |  | 3021 | 1.9199 |  |
| C. | CD16 Mono | 165 | Day 14 |  | 4634 | 3.5606 |  |
| D. | CD16 Mono | 25 | Day 14 |  | 5605 | 0.4460 |  |
| E. | CD16 Mono | 64 | Day 14 | 0.4375 | 5377 | 1.1903 | 0.3125 |
| A. | CD4 Memory | 1087 | Day 0 |  | 4074 | 26.6814 |  |
| B. | CD4 Memory | 1814 | Day 0 |  | 5501 | 32.9758 |  |
| C. | CD4 Memory | 1865 | Day 0 |  | 10930 | 17.0631 |  |
| D. | CD4 Memory | 2808 | Day 0 |  | 8527 | 32.9307 |  |
| E. | CD4 Memory | 994 | Day 0 |  | 5396 | 18.4211 |  |
| A. | CD4 Memory | 45 | Day 14 |  | 278 | 16.1871 |  |
| B. | CD4 Memory | 915 | Day 14 |  | 3021 | 30.2880 |  |
| C. | CD4 Memory | 778 | Day 14 |  | 4634 | 16.7890 |  |
| D. | CD4 Memory | 1422 | Day 14 |  | 5605 | 25.3702 |  |
| E. | CD4 Memory | 903 | Day 14 | 0.0625 | 5377 | 16.7938 | 0.0625 |
| A. | CD4 Naive T | 1245 | Day 0 |  | 4074 | 30.5596 |  |
| B. | CD4 Naive T | 1260 | Day 0 |  | 5501 | 22.9049 |  |
| C. | CD4 Naive T | 1686 | Day 0 |  | 10930 | 15.4254 |  |
| D. | CD4 Naive T | 1788 | Day 0 |  | 8527 | 20.9687 |  |
| E. | CD4 Naive T | 1227 | Day 0 |  | 5396 | 22.7391 |  |
| A. | CD4 Naive T | 48 | Day 14 |  | 278 | 17.2862 |  |
| B. | CD4 Naive T | 609 | Day 14 |  | 3021 | 20.1589 |  |
| C. | CD4 Naive T | 887 | Day 14 |  | 4634 | 19.1411 |  |
| D. | CD4 Naive T | 842 | Day 14 |  | 5605 | 15.0223 |  |
| E. | CD4 Naive T | 1243 | Day 14 | 0.1250 | 5377 | 23.1170 | 0.4375 |
| A. | CD8 T | 459 | Day 0 |  | 4074 | 11.2666 |  |
| B. | CD8 T | 542 | Day 0 |  | 5501 | 9.8528 |  |
| C. | CD8 T | 3107 | Day 0 |  | 10930 | 28.4263 |  |
| D. | CD8 T | 1193 | Day 0 |  | 8527 | 13.9909 |  |
| E. | CD8 T | 463 | Day 0 |  | 5396 | 8.5804 |  |
| A. | CD8 T | 15 | Day 14 |  | 278 | 5.3957 |  |
| B. | CD8 T | 287 | Day 14 |  | 3021 | 9.5002 |  |
| C. | CD8 T | 967 | Day 14 |  | 4634 | 20.8675 |  |
| D. | CD8 T | 846 | Day 14 |  | 5605 | 15.0937 |  |
| E. | CD8 T | 390 | Day 14 | 0.0625 | 5377 | 7.2531 | 0.1875 |
| A. | DC | 15 | Day 0 |  | 4074 | 0.3682 |  |
| B. | DC | 45 | Day 0 |  | 5501 | 0.8180 |  |
| C. | DC | 19 | Day 0 |  | 10930 | 0.1738 |  |
| D. | DC | 15 | Day 0 |  | 8527 | 0.1759 |  |
| E. | DC | 44 | Day 0 |  | 5396 | 0.8154 |  |
| A. | DC | 3 | Day 14 |  | 278 | 1.0791 |  |
| B. | DC | 15 | Day 14 |  | 3021 | 0.4965 |  |
| C. | DC | 11 | Day 14 |  | 4634 | 0.2374 |  |
| D. | DC | 12 | Day 14 |  | 5605 | 0.2141 |  |
| E. | DC | 61 | Day 14 | 0.4375 | 5377 | 1.1345 | 0.4375 |
| A. | Eryth | 8 | Day 0 |  | 4074 | 0.1964 |  |
| B. | Eryth | 3 | Day 0 |  | 5501 | 0.0545 |  |
| C. | Eryth | 12 | Day 0 |  | 10930 | 0.1098 |  |
| D. | Eryth | 238 | Day 0 |  | 8527 | 2.7911 |  |
| E. | Eryth | 127 | Day 0 |  | 5396 | 2.3536 |  |
| A. | Eryth | 1 | Day 14 |  | 278 | 0.3597 |  |
| B. | Eryth | 3 | Day 14 |  | 3021 | 0.0993 |  |
| C. | Eryth | 15 | Day 14 |  | 4634 | 0.3237 |  |
| D. | Eryth | 261 | Day 14 |  | 5605 | 4.6566 |  |
| E. | Eryth | 76 | Day 14 | 0.8551 | 5377 | 1.4134 | 0.4375 |
| A. | MK | 34 | Day 0 |  | 4074 | 0.8346 |  |
| B. | MK | 5 | Day 0 |  | 5501 | 0.0909 |  |
| C. | MK | 129 | Day 0 |  | 10930 | 1.1802 |  |
| D. | MK | 95 | Day 0 |  | 8527 | 1.1141 |  |
| E. | MK | 25 | Day 0 |  | 5396 | 0.4633 |  |
| A. | MK | 1 | Day 14 |  | 278 | 0.3597 |  |
| B. | MK | 11 | Day 14 |  | 3021 | 0.3641 |  |
| C. | MK | 50 | Day 14 |  | 4634 | 1.0790 |  |
| D. | MK | 59 | Day 14 |  | 5605 | 1.0526 |  |
| E. | MK | 9 | Day 14 | 0.1250 | 5377 | 0.1674 | 0.3125 |
| A. | NK | 16 | Day 0 |  | 4074 | 0.3927 |  |
| B. | NK | 18 | Day 0 |  | 5501 | 0.3272 |  |
| C. | NK | 18 | Day 0 |  | 10930 | 0.1647 |  |
| D. | NK | 16 | Day 0 |  | 8527 | 0.1876 |  |
| E. | NK | 10 | Day 0 |  | 5396 | 0.1853 |  |
| A. | NK | 1 | Day 14 |  | 278 | 0.3597 |  |
| B. | NK | 20 | Day 14 |  | 3021 | 0.6620 |  |
| C. | NK | 126 | Day 14 |  | 4634 | 2.7190 |  |
| D. | NK | 31 | Day 14 |  | 5605 | 0.5531 |  |
| E. | NK | 14 | Day 14 | 0.3430 | 5377 | 0.2604 | 0.1250 |
| A. | pDC | 2 | Day 0 |  | 4074 | 0.0491 |  |
| B. | pDC | 14 | Day 0 |  | 5501 | 0.2545 |  |
| C. | pDC | 7 | Day 0 |  | 10930 | 0.0640 |  |
| D. | pDC | 6 | Day 0 |  | 8527 | 0.0704 |  |
| E. | pDC | 20 | Day 0 |  | 5396 | 0.3706 |  |
| A. | pDC | 3 | Day 14 |  | 278 | 1.0791 |  |
| B. | pDC | 29 | Day 14 |  | 3021 | 0.9599 |  |
| C. | pDC | 9 | Day 14 |  | 4634 | 0.1942 |  |
| D. | pDC | 12 | Day 14 |  | 5605 | 0.2141 |  |
| E. | pDC | 21 | Day 14 | 0.0579 | 5377 | 0.3906 | 0.0625 |
| A. | T Activated | 206 | Day 0 |  | 4074 | 5.0565 |  |
| B. | T Activated | 1155 | Day 0 |  | 5501 | 20.9962 |  |
| C. | T Activated | 2403 | Day 0 |  | 10930 | 21.9854 |  |
| D. | T Activated | 1051 | Day 0 |  | 8527 | 12.3256 |  |
| E. | T Activated | 730 | Day 0 |  | 5396 | 13.5285 |  |
| A. | T Activated | 13 | Day 14 |  | 278 | 4.6763 |  |
| B. | T Activated | 800 | Day 14 |  | 3021 | 26.4813 |  |
| C. | T Activated | 968 | Day 14 |  | 4634 | 20.8891 |  |
| D. | T Activated | 974 | Day 14 |  | 5605 | 17.3773 |  |
| E. | T Activated | 776 | Day 14 | 0.1250 | 5377 | 14.4318 | 0.4375 |

**Table S5.** Splenic Ultrasound in Healthy Participants: Subject Features. Characteristics and demographic data of 20 enrolled subjects for study of biological effects of ultrasound of the spleen. Ten subjects received full-powered ultrasound treatment at the spleen hilum and another ten subjects were sham controls with the ultrasound device turned off. One of the subjects from the stimulated group was excluded due the selection of the wrong ultrasound imaging protocol by the clinician. One of the subjects from the control group was excluded because their baseline glucose measurement was <50 mg/dL that is considered hypoglycemic.

| Subject | 1 | 2 | 3 | 4 | 5 | 6 | 7 | 8 | 9 | 10 |
| --- | --- | --- | --- | --- | --- | --- | --- | --- | --- | --- |
| Group | Stimulated | Stimulated | Stimulated | Stimulated | Stimulated | Stimulated | Stimulated | Stimulated | Stimulated | Stimulated |
| Age | 35 | 39 | 29 | 39 | 44 | 30 | 28 | 35 | 32 | 42 |
| Gender | Female | Female | Male | Female | Female | Female | Male | Female | Female | Female |
| Race | Asian | Asian | More Than One Race | Other | Black or African American | More Than One Race | Other | Other | White or Caucasian | White or Caucasian |
| Ethnicity | Not Hispanic or Latino | Not Hispanic or Latino | Not Hispanic or Latino | Hispanic or Latino | Not Hispanic or Latino | Hispanic or Latino | Not Hispanic or Latino | Hispanic or Latino | Hispanic or Latino | Not Hispanic or Latino |
| Height (cm) | 177 | 165 | 188 | 159 | 164 | 168 | 195 | 160 | 156 | 168 |
| Weight (kg) | 64 | 58.5 | 94.6 | 69.8 | 64.5 | 67 | 79.1 | 55.8 | 58.7 | 70 |
| Body Mass Index (kg/m2) | 20.4 | 21.5 | 26.8 | 27.6 | 24 | 23.7 | 20.8 | 21.8 | 24.1 | 24.8 |
| Medicines & Supplements |  |  | Multi Vitamin | Multi Vitamin | Multi Vitamin |  |  |  |  |  |
| Conditions |  |  | Hemo-phagocytic Lymphohistiocytosis 14 years ago |  |  |  |  |  | Gestational diabetes in prior 3 years. Resolved after delivery. | Mitral Value Prolapse, Current - No treatment needed |
| Exclusion Reason | Excluded : incorrect imaging protocol applied |  |  |  |  |  |  |  |  |  |

| Subject | 11 | 12 | 13 | 14 | 15 | 16 | 17 | 18 | 19 | 20 |
| --- | --- | --- | --- | --- | --- | --- | --- | --- | --- | --- |
| Group | Control | Control | Control | Control | Control | Control | Control | Control | Control | Control |
| Age | 22 | 27 | 32 | 18 | 36 | 33 | 27 | 40 | 28 | 28 |
| Gender | Female | Male | Female | Male | Male | Female | Female | Male | Male | Male |
| Race | White or Caucasian | Asian | Black or African American | Asian | Asian | Asian | Asian | Asian | Asian | Other |
| Ethnicity | Not Hispanic or Latino | Not Hispanic or Latino | Not Hispanic or Latino | Not Hispanic or Latino | Not Hispanic or Latino | Not Hispanic or Latino | Not Hispanic or Latino | Not Hispanic or Latino | Not Hispanic or Latino | Hispanic or Latino |
| Height (cm) | 162 | 176 | 173 | 183 | 176 | 161 | 155 | 166 | 172.25 | 180 |
| Weight (kg) | 73.9 | 72 | 72.9 | 72.3 | 80.9 | 58.2 | 61.1 | 67 | 81.5 | 75.4 |
| Body Mass Index (kg/m2) | 28.2 | 23.2 | 24.4 | 21.6 | 26.1 | 22.5 | 25.4 | 24.3 | 27.5 | 23.3 |
| Medicines & Supplements |  | Vitamin D, EOD 5000 IU, Promote Health |  |  | Multi Vitamin, Fish Oil | Multi Vitamin |  |  | Pantoperazole 20 mg daily, acid reflux | Multi Vitamin |
| Conditions | Acid Reflux |  |  |  | Jaundice at 7 years of age, resolved |  |  |  |  |  |
| Exclusion Reason |  |  |  |  |  |  |  |  | Excluded : glucose <50 mg/dL at baseline |  |

